# Menopausal hormone therapy and the female brain: leveraging neuroimaging and prescription registry data from the UK Biobank cohort

**DOI:** 10.1101/2024.04.08.24305450

**Authors:** Claudia Barth, Liisa A.M. Galea, Emily G. Jacobs, Bonnie H. Lee, Lars T. Westlye, Ann-Marie G. de Lange

**Author notes:** **Corresponding author:** Claudia Barth, PhD.

## Abstract

**Background and Objectives:** Menopausal hormone therapy (MHT) is generally thought to be neuroprotective, yet results have been inconsistent. Here, we present a comprehensive study of MHT use and brain characteristics in middle-to older aged females from the UK Biobank, assessing detailed MHT data, APOE ε4 genotype, and tissue-specific gray (GM) and white matter (WM) brain age gap (BAG), as well as hippocampal and white matter hyperintensity (WMH) volumes.

**Methods:** A total of 19,846 females with magnetic resonance imaging data were included (current-users = 1,153, 60.1 ± 6.8 years; past-users = 6,681, 67.5 ± 6.2 years; never-users = 12,012, mean age 61.6 ± 7.1 years). For a sub-sample (n = 538), MHT prescription data was extracted from primary care records. Brain measures were derived from T1-, T2- and diffusion-weighted images. We fitted regression models to test for associations between the brain measures and MHT variables including user status, age at initiation, dosage and duration, formulation, route of administration, and type (i.e., bioidentical vs synthetic), as well as active ingredient (e.g., estradiol hemihydrate). We further tested for differences in brain measures among MHT users with and without a history of hysterectomy ± bilateral oophorectomy and examined associations by APOE ε4 status.

**Results:** We found significantly higher GM and WM BAG (i.e., older brain age relative to chronological age) as well as smaller left and right hippocampus volumes in current MHT users, not past users, compared to never-users. Effects were modest, with the largest effect size indicating a group difference of 0.77 years (∼9 months) for GM BAG. Among MHT users, we found no significant associations between age at MHT initiation and brain measures. Longer duration of use and older age at last use post menopause was associated with higher GM and WM BAG, larger WMH volume, and smaller left and right hippocampal volumes. MHT users with a history of hysterectomy ± bilateral oophorectomy showed *lower* GM BAG relative to MHT users without such history. Although we found smaller hippocampus volumes in carriers of two APOE ε4 alleles compared to non-carriers, we found no interactions with MHT variables. In the sub-sample with prescription data, we found no significant associations between detailed MHT variables and brain measures after adjusting for multiple comparisons.

**Discussion:** Our results indicate that population-level associations between MHT use, and female brain health might vary depending on duration of use and past surgical history. Future research is crucial to establish causality, dissect interactions between menopause-related neurological changes and MHT use, and determine individual-level implications to advance precision medicine in female health care.

## Introduction

Ovarian hormones such as estrogens and progesterone fluctuate across the female lifespan with natural declines occurring at menopause, typically between the ages 45 and 55. The cessation of ovarian function during the menopausal transition has been linked to an array of neural changes ^1^, including a decline in brain glucose metabolism ^2^, reductions in gray matter (GM) and white matter (WM) volume ^3–6^, and increased amyloid-beta deposition ^7^ as well as WM lesions ^8^. In combination with other risk factors, these neural changes might foster the emergence of neurodegenerative diseases such as late-onset Alzheimer’s disease (AD), which is more often diagnosed in females relative to similarly aged males, with greater cognitive decline and neuropathological burden ^9, 10^.

Menopausal hormone therapy (MHT) is commonly prescribed to minimize vasomotor symptoms occurring during the menopausal transition, and is generally thought to be neuroprotective, with a propensity to reduce the risk for AD ^11–14^ and improve cognition later in life ^15^. However, study results are equivocal ^16^, reporting both positive ^17–19^ and negative outcomes ^20–22^. A 2021 study reported that reproductive history events signaling more estrogens exposure, including MHT use, were associated with greater gray matter volume in middle-aged females ^17^, in line with other neuroimaging studies suggesting a protective effect of MHT on GM, WM, and ventricle size ^18, 19^. Conversely, MHT use has also been associated with greater atrophy ^23^ and higher rates of ventricular expansion ^20^ in menopausal females. Similarly, in our previous UK Biobank study of ∼16,000 females ^21^, we found positive associations between MHT use and older GM brain age, albeit with small effect sizes.

Besides mixed results in observational studies, randomized controlled trials (RCTs) such as the Women’s Health Initiative Memory Study (WHIMS) suggest an increased risk of dementia and cognitive decline with MHT use. In detail, WHIMS found negative effects of prolonged oral administration of both conjugated equine estrogen (CEE) alone ^24^ or in combination with medroxyprogesterone acetate (MPA, synthetic progestin) ^25, 26^ among females aged 65 years or older. Similarly, the Heart and Estrogen/Progestin Replacement Study (HERS) showed an association between 4 years of CEE + MPA treatment and lower cognitive performance in older postmenopausal females (71 ± 6 years) ^27^. In contrast, administering oral CEE or transdermal estradiol plus micronized progesterone in recently postmenopausal females did not alter cognition in the Kronos Early Estrogen Prevention Study (KEEPS) ^28^. These mixed findings raise the question of whether a combination of timing, formulation, and route of administration might play a crucial role in the effectiveness of MHT.

According to the ‘*critical window hypothesis*’, MHT might be neuroprotective if it is initiated close to menopause ^29^. For example, MHT initiation during perimenopause has been associated with improved memory and hippocampal function later in life ^15^. Although emerging evidence supports this hypothesis ^30, 31^, oral CEE use in combination with MPA has been found to increase the risk for memory decline regardless of timing ^26, 29, 32^. Similarly, systemic MHT (i.e. oral and transdermal use) has been associated with a 9-17% increased risk of AD in a Finish nationwide case-control study, independent of MHT formulation and timing ^33^. However, vaginal estradiol use did not show such risk, indicating differential effects of route of administration ^33^. Vaginal as well as transdermal MHT formulations are mainly composed of estradiol, and CEE is a blend of compounds, mostly estrone ^34^. Both estrogens have different affinities to bind to estrogen receptors (ER): relative to estradiol, estrone is approximately 2/3 the affinity to ER-alpha, and about 1/3 to ER-beta ^35^, and estrone is present at higher levels post-menopause than estradiol. Furthermore, contrary to oral use, vaginal and transdermal MHT formulations bypass hepatic metabolism, resulting in steady-state concentration of estradiol (estrone:estradiol ratio of 1:1) ^34^. Hence, vaginal, or transdermal estradiol-based MHT formulations might be more effective than oral estrone-based types. For instance, Gleason and colleagues found that females exposed to oral CEE exhibited poorer memory performance than either MHT-naïve or estradiol-exposed individuals ^36^. In addition to estrogens, progestins are commonly added in non-hysterectomized females, and, like estrogens based MHT, progestins can be administered in different forms, e.g., norethisterone acetate (synthetic progestin), micronized progesterone (bioidentical), or MPA (synthetic progestin). These progestin forms have been linked to different side-effect profiles ^37^ and can antagonize the effects of estrogens in MHT ^38^.

In addition to the impact of MHT timing, formulation, and route of administration, effects of MHT on the female brain might be modulated by apolipoprotein ε type 4 (APOE ε4) genotype. Carried by 14% of the world’s population, the APOE ε4 allele is a known dose-dependent risk factor for late-onset AD. Yaffe and colleagues found that among non-carriers, current MHT use lowered the risk of cognitive impairment by almost half compared to never-users, while there was no such effect among carriers ^39^. Results from the Nurses’ Health Study found that MHT use was associated with *worse* rates of decline in general cognition, especially among females with an APOE ε4 allele ^40^. Conversely, we found no significant interactions between APOE ε4 genotype and MHT use on cognition in a 2023 UK Biobank study ^41^. We did however observe that earlier MHT initiation was linked to younger GM brain age, albeit weakly, only in APOE ε4 carriers ^21^. Moreover, a 2024 UK Biobank study showed smaller brain volumes in MHT users compared to never-users, specifically among females with the APOE ε4/ε4 genotype ^42^. Yet, the potential of APOE ε4 to modulate effects of MHT dosage, administration, and formulation on the female brain is largely unexplored.

In summary, emerging evidence suggests differential effects of MHT formulation, age at initiation, route of administration, and genotype on female brain and cognition. However, studies aiming to disentangle the effects of different MHT regimes are largely missing ^43^. In this observational study, we investigated associations between MHT variables, different MHT regimes, APOE ε4 status, and brain measures in middle to older-aged females from the UK Biobank cohort. MHT variables included user status (i.e. current users, past users, never users), age at initiation, dosage and duration, formulation, route of administration, and type (i.e., bioidentical vs synthetic) as well as active ingredient (e.g., estradiol hemihydrate). MHT regimes based on prescription data were extracted from primary care records (general practice). Brain measures included GM and WM brain age gap based on brain age prediction, hippocampal volumes, and total WM hyperintensity volume as proxy of vascular disease. These measures were chosen as they have been linked to both chronological and endocrine aging as well as MHT use in our studies^21, 42, 44, 45^.

## Methods

### Sample characteristics

The sample was drawn from the UK Biobank cohort (www.ukbiobank.ac.uk). Females with diffusion-, and T1-weighted magnetic resonance imaging (MRI) data and complete data on demographic factors, lifestyle factors, and BMI from the MRI assessment time point were included, yielding a sample of 20,325. Out of this sample, a total of 19,846 participants had complete data related to MHT user status (**Table 1-2**), and 14,693 participants had complete data on APOE ε4 status. These samples provided the basis for the general MHT-use analyses. Among the MHT users, a subsample of 538 participants had complete MHT-related prescription data and MRI data (**Table 3**, n = 521 with data on APOE ε4 status).

**Table 1.** Sample demographics of menopausal hormone therapy (MHT) never-, current, and past-users in the whole sample.

|  | MHT User Status |  |  | p-value |  |  |
| --- | --- | --- | --- | --- | --- | --- |
|  | Never | Current | Past | Never vs Current | Never vs Past | Current vs Past |
| **N** | **12,012** | **1,153** | **6,681** |  |  |  |
| **Age\*** | 61.6 ± 7.1 | 60.1 ± 6.8 | 67.5 ± 6.2 | **<0.001** | **<0.001** | **<0.001** |
| **Ethnic Background, N (%)** |  |  |  | 0.084 | **<0.001** | 0.824 |
| White | 11,590 (96.6) | 1,128 (98.0) | 6,555 (98.2) |  |  |  |
| Asian | 103 (0.9) | 5 (0.4) | 27 (0.4) |  |  |  |
| Black | 96 (0.8) | 4 (0.3) | 23 (0.3) |  |  |  |
| Chinese | 54 (0.5) | 1 (0.1) | 12 (0.2) |  |  |  |
| Mixed | 81 (0.7) | 5 (0.4) | 27 (0.4) |  |  |  |
| Other ethnic group | 73 (0.6) | 8 (0.7) | 28 (0.4) |  |  |  |
| **Education, N (%)** |  |  |  | 0.260 | **<0.001** | **<0.001** |
| College/University degree | 6,123 (51.0) | 606 (52.6) | 2,813 (42.1) |  |  |  |
| O levels/GCSEs or equivalent | 2,234 (18.6) | 203 (17.6) | 1,447 (21.7) |  |  |  |
| A levels/AS levels or equivalent | 1,656 (13.8) | 135 (11.7) | 758 (11.3) |  |  |  |
| CSEs or equivalent | 471 (3.9) | 52 (4.5) | 247 (3.7) |  |  |  |
| NVQ/HND/HNC or equivalent | 414 (3.4) | 38 (3.3) | 279 (4.2) |  |  |  |
| Other professional qualifications | 602 (5.0) | 70 (6.1) | 523 (7.8) |  |  |  |
| None of the above | 512 (4.3) | 49 (4.2) | 613 (9.2) |  |  |  |
| **Lifestyle score** | 1.7 ± 1.2 | 1.8 ± 1.3 | 1.7 ± 1.2 | **0.009** | 0.076 | 0.089 |
| **BMI\* (m<sup>2</sup>/kg)** | 26.0 ± 4.8 | 25.5 ± 4.4 | 26.2 ± 4.6 | **0.003** | **0.003** | **<0.001** |
| **Menopausal Status, N (%)** |  |  |  | **<0.001** | **<0.001** | **<0.001** |
| No | 962 (8.0) | 45 (3.9) | 37 (0.6) |  |  |  |
| Yes | 9,905 (82.5) | 769 (66.7) | 5,528 (82.8) |  |  |  |
| Not sure – had a hysterectomy | 545 (4.5) | 247 (21.4) | 1,057 (15.8) |  |  |  |
| Not sure – other reason | 600 (5.0) | 92 (8.0) | 58 (0.9) |  |  |  |
| **Oophorectomy, yes, N (%)** | 443 (3.7) | 224 (19.4) | 1172 (17.7) | **<0.001** | **<0.001** | 0.159 |
| **Hysterectomy, yes, N (%)** | 518 (4.5) | 117 (12.5) | 1083 (18.8) | **<0.001** | **<0.001** | **<0.001** |
| **APOE ε4 status, carrier, N (%)** | 3134 (27.5) | 284 (26.2) | 1557 (24.7) | 0.398 | **<0.001** | 0.278 |
| **APOE ε4, allele, N (%)** |  |  |  | 0.649 | **<0.001** | 0.460 |
| non-carrier | 8264 (72.5) | 798 (73.8) | 4759 (75.3) |  |  |  |
| ε3/ε4 | 2853 (25.0) | 257 (23.8) | 1424 (22.5) |  |  |  |
| ε4/ ε4 | 281 (2.5) | 27 (2.5) | 133 (2.1) |  |  |  |
| **Age started MHT\*** |  | 49.8 ± 6.5 | 47.9 ± 5.6 |  |  | **<0.001** |
| **Age last used MHT\*** |  | 60.1 ± 6.8 | 53.9 ± 6.1 |  |  | **<0.001** |
| **Duration of MHT use\*** |  | 10.3 ± 8.6 | 6.0 ± 5.6 |  |  | **<0.001** |

\* Mean ± Standard Deviation. Age is given in years. Abbreviations: N, sample size; GCSE, General Certificate of Secondary Education; CSE, Certificate of Secondary Education; NVQ, National Vocational Qualification; BMI, body mass index; APOE, apolipoprotein. Significant differences between groups based on t/χ<sup>2</sup> tests are highlighted in bold.

**Table 2.** Sample demographics of menopausal hormone therapy (MHT) users with and without a history of hysterectomy +/-bilateral oophorectomy in the whole sample.

| N | MHT Users |  |  | p-value |  |  |
| --- | --- | --- | --- | --- | --- | --- |
|  | No Surgery<br>**5,510** | Hysterectomy<br>**544** | Oophorectomy<br>**1,407** | No vs<br>Hyster | No vs<br>Oopho | Hyster vs<br>Oopho |
| **Age\*** | 66.1 ± 6.9 | 69.0 ± 5.6 | 66.7 ± 6.7 | **<0.001** | **0.003** | **<0.001** |
| **Ethnic Background, N (%)** |  |  |  | 0.710 | 0.272 | 0.569 |
| White | 5,408 (98.3) | 535 (98.5) | 1,370 (97.4) |  |  |  |
| Asian | 24 (0.4) | 2 (0.4) | 8 (0.6) |  |  |  |
| Black | 15 (0.3) | 3 (0.6) | 8 (0.6) |  |  |  |
| Chinese | 9 (0.2) | 0 (0.0) | 4 (0.3) |  |  |  |
| Mixed | 22 (0.4) | 2 (0.4) | 6 (0.4) |  |  |  |
| Other ethnic group | 23 (0.4) | 1 (0.2) | 10 (0.7) |  |  |  |
| **Education, N (%)** |  |  |  | **<0.001** | **<0.001** | 0.197 |
| College/University degree | 2,559 (46.4) | 184 (33.8) | 557 (39.6) |  |  |  |
| O levels/GCSEs or equivalent | 1,099 (19.9) | 132 (24.3) | 323 (23.0) |  |  |  |
| A levels/AS levels or equivalent | 631 (11.5) | 66 (12.1) | 156 (11.1) |  |  |  |
| CSEs or equivalent | 206 (3.7) | 18 (3.3) | 54 (3.8) |  |  |  |
| NVQ/HND/HNC or equivalent | 209 (3.8) | 24 (4.4) | 63 (4.5) |  |  |  |
| Other professional qualifications | 387 (7.0) | 60 (11.0) | 114 (8.1) |  |  |  |
| None of the above | 419 (7.6) | 60 (11.0) | 140 (10.0) |  |  |  |
| **Lifestyle score\*** | 1.7 ± 1.2 | 1.6 ± 1.2 | 1.8 ± 1.2 | 0.323 | 0.066 | 0.044 |
| **BMI (kg/m<sup>2</sup>) \*** | 25.9 ± 4.50 | 26.1 ± 4.2 | 26.7 ± 4.7 | 0.260 | **<0.001** | **0.011** |
| **Menopausal Status, N (%)** |  |  |  | **<0.001** | **<0.001** | **<0.001** |
| No | 81 (1.5) | 0 (0.0) | 1 (0.1) |  |  |  |
| Yes | 5,290 (96.0) | 444 (81.6) | 615 (43.7) |  |  |  |
| Not sure – had a hysterectomy | 0 (0.0) | 99 (18.2) | 781 (55.5) |  |  |  |
| Not sure – other reason | 139 (2.5) | 1 (0.2) | 10 (0.7) |  |  |  |
| **Hysterectomy, yes, N (%)** |  |  | 666 (89.4) |  |  |  |
| **APOE ε4 status, carrier, N (%)** | 1317 (25.3) | 113 (21.9) | 345 (25.9) | 0.100 | 0.678 | 0.085 |
| **APOE ε4, allele, N (%)** |  |  |  | 0.095 | 0.427 | 0.161 |
| non-carrier | 3881 (74.7) | 402 (78.1) | 985 (74.1) |  |  |  |
| ε3/ε4 | 1194 (23.0) | 107 (20.8) | 320 (24.1) |  |  |  |
| ε4/ ε4 | 123 (2.4) | 6 (1.2) | 25 (1.9) |  |  |  |
| **Age at menopause\*** | 50.0 ± 5.1 | 43.1 ± 6.8 | 46.9 ± 6.3 | **<0.001** | **<0.001** | **<0.001** |
| **Age started MHT\*** | 49.2 ± 5.3 | 46.9 ± 5.7 | 45.6 ± 6.1 | **<0.001** | **<0.001** | **<0.001** |
| **Age last used MHT\*** | 54.7 ± 6.1 | 55.0 ± 7.4 | 55.2 ± 7.4 | 0.312 | **0.014** | 0.646 |
| **Duration of MHT use\*** | 5.5 ± 5.4 | 8.1 ± 6.6 | 9.6 ± 7.8 | **<0.001** | **<0.001** | **<0.001** |
| **Age MHT rel Age Menopause** |  |  |  | **<0.001** | **<0.001** | **<0.001** |
| Same age | 1177 (24.7) | 89 (19.7) | 303 (46.2) |  |  |  |
| After | 1659 (34.8) | 283 (62.7) | 170 (25.9) |  |  |  |
| Before | 1931 (40.5) | 79 (17.5) | 183 (27.9) |  |  |  |
| **Age at oophorectomy\*** |  |  | 47.7 ± 8.2 |  |  |  |
| **Age at hysterectomy\*** |  | 44.5 ± 10.0 | 46.9 ± 7.9 |  |  | **<0.001** |
| **Age Oopho rel Age Menopause** |  |  |  |  |  |  |
| Same age |  |  | 301 (47.8) |  |  |  |
| After |  |  | 271 (43.0) |  |  |  |
| Before |  |  | 58 (9.2) |  |  |  |
| **Age Oopho rel Age MHT** |  |  |  |  |  |  |
| Same age |  |  | 740 (59.0) |  |  |  |
| After |  |  | 367 (29.2) |  |  |  |
| Before |  |  | 148 (11.8) |  |  |  |
| **Age Hyster rel Age Menopause** |  |  |  |  |  | **<0.001** |
| Same age |  | 278 (61.6) | 324 (54.8) |  |  |  |
| After |  | 110 (24.4) | 214 (36.2) |  |  |  |
| Before |  | 63 (14.0) | 53 (9.0) |  |  |  |
| **Age Hyster rel Age MHT** |  |  |  |  |  | **<0.001** |
| Same age |  | 80 (17.2) | 746 (61.2) |  |  |  |
| After |  | 111 (23.9) | 296 (24.3) |  |  |  |
| Before |  | 274 (58.9) | 177 (14.5) |  |  |  |

\* Mean ± Standard Deviation. Age is given in years. Hysterectomy/hyster included females without bilateral oophorectomy; Oophorectomy/Oopho constitutes bilateral oophorectomy (+/- hysterectomy; no hysterectomy n = X, with hysterectomy n = Y). Abbreviation: N, sample size; GCSE, General Certificate of Secondary Education; CSE, Certificate of Secondary Education; NVQ, National Vocational Qualification; BMI, body mass index; APOE, apolipoprotein. Significant differences between groups based on t/χ<sup>2</sup> tests are highlighted in bold.

**Table 3.**
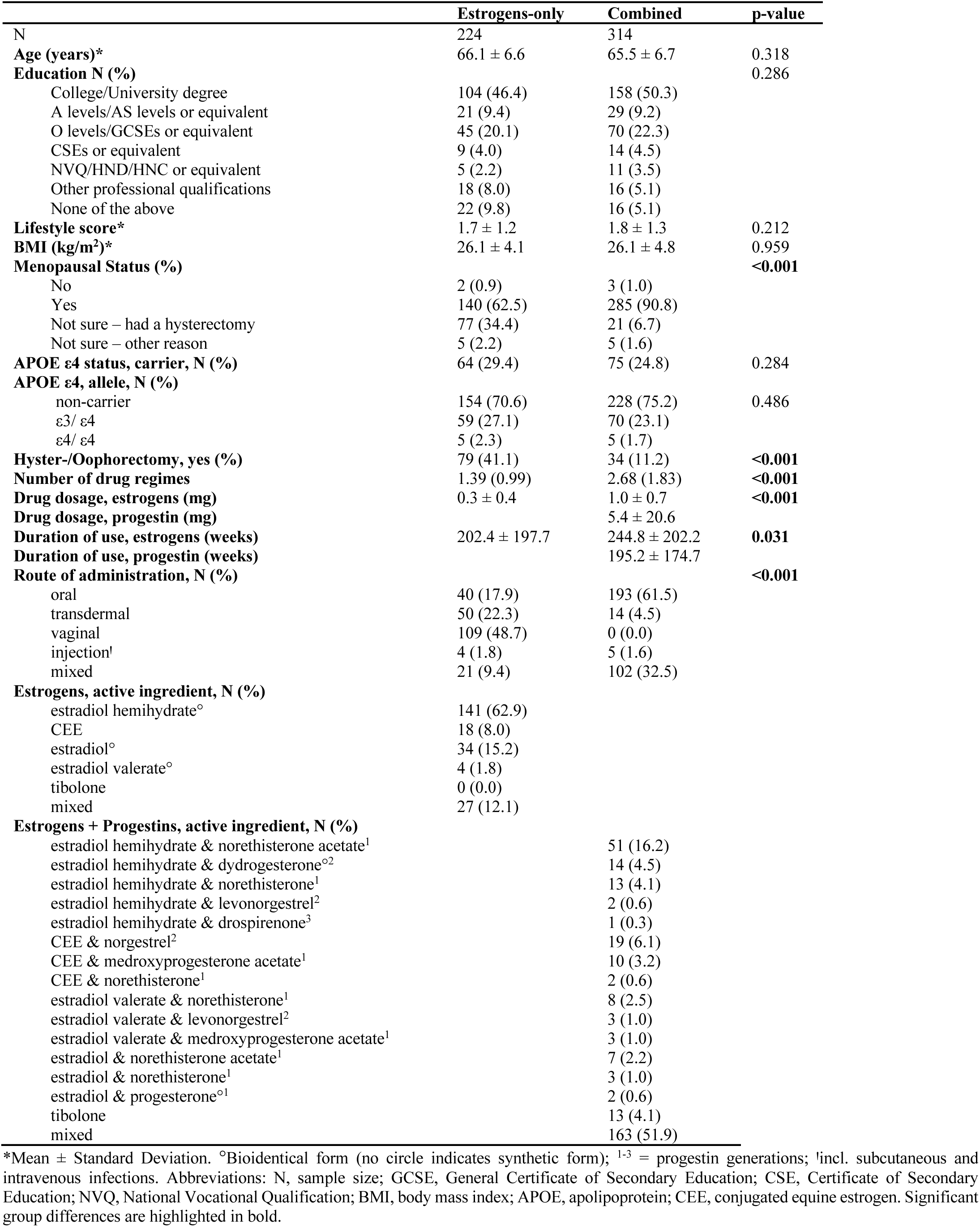
Sample demographics of menopausal hormone therapy (MHT) users with prescription data, stratified by estrogen only MHT or combined MHT use.

|  | Estrogens-only | Combined | p-value |
| --- | --- | --- | --- |
| N | 224 | 314 |  |
| Age (years)* | 66.1 ± 6.6 | 65.5 ± 6.7 | 0.318 |
| Education N (%) |  |  | 0.286 |
| College/University degree | 104 (46.4) | 158 (50.3) |  |
| A levels/AS levels or equivalent | 21 (9.4) | 29 (9.2) |  |
| O levels/GCSEs or equivalent | 45 (20.1) | 70 (22.3) |  |
| CSEs or equivalent | 9 (4.0) | 14 (4.5) |  |
| NVQ/HND/HNC or equivalent | 5 (2.2) | 11 (3.5) |  |
| Other professional qualifications | 18 (8.0) | 16 (5.1) |  |
| None of the above | 22 (9.8) | 16 (5.1) |  |
| Lifestyle score* | 1.7 ± 1.2 | 1.8 ± 1.3 | 0.212 |
| BMI (kg/m<sup>2</sup>)* | 26.1 ± 4.1 | 26.1 ± 4.8 | 0.959 |
| Menopausal Status (%) |  |  | **<0.001** |
| No | 2 (0.9) | 3 (1.0) |  |
| Yes | 140 (62.5) | 285 (90.8) |  |
| Not sure – had a hysterectomy | 77 (34.4) | 21 (6.7) |  |
| Not sure – other reason | 5 (2.2) | 5 (1.6) |  |
| APOE ε4 status, carrier, N (%) | 64 (29.4) | 75 (24.8) | 0.284 |
| APOE ε4, allele, N (%) |  |  |  |
| non-carrier | 154 (70.6) | 228 (75.2) | 0.486 |
| ε3/ ε4 | 59 (27.1) | 70 (23.1) |  |
| ε4/ ε4 | 5 (2.3) | 5 (1.7) |  |
| Hyster-/Oophorectomy, yes (%) | 79 (41.1) | 34 (11.2) | **<0.001** |
| Number of drug regimes | 1.39 (0.99) | 2.68 (1.83) | **<0.001** |
| Drug dosage, estrogens (mg) | 0.3 ± 0.4 | 1.0 ± 0.7 | **<0.001** |
| Drug dosage, progestin (mg) |  | 5.4 ± 20.6 |  |
| Duration of use, estrogens (weeks) | 202.4 ± 197.7 | 244.8 ± 202.2 | **0.031** |
| Duration of use, progestin (weeks) |  | 195.2 ± 174.7 |  |
| Route of administration, N (%) |  |  | **<0.001** |
| oral | 40 (17.9) | 193 (61.5) |  |
| transdermal | 50 (22.3) | 14 (4.5) |  |
| vaginal | 109 (48.7) | 0 (0.0) |  |
| injection<sup>l</sup> | 4 (1.8) | 5 (1.6) |  |
| mixed | 21 (9.4) | 102 (32.5) |  |
| Estrogens, active ingredient, N (%) |  |  |  |
| estradiol hemihydrate<sup>o</sup> | 141 (62.9) |  |  |
| CEE | 18 (8.0) |  |  |
| estradiol<sup>o</sup> | 34 (15.2) |  |  |
| estradiol valerate<sup>o</sup> | 4 (1.8) |  |  |
| tibolone | 0 (0.0) |  |  |
| mixed | 27 (12.1) |  |  |
| Estrogens + Progestins, active ingredient, N (%) |  |  |  |
| estradiol hemihydrate & norethisterone acetate<sup>1</sup> |  | 51 (16.2) |  |
| estradiol hemihydrate & dydrogesterone<sup>o2</sup> |  | 14 (4.5) |  |
| estradiol hemihydrate & norethisterone<sup>1</sup> |  | 13 (4.1) |  |
| estradiol hemihydrate & levonorgestrel<sup>2</sup> |  | 2 (0.6) |  |
| estradiol hemihydrate & drospirenone<sup>3</sup> |  | 1 (0.3) |  |
| CEE & norgestrel<sup>2</sup> |  | 19 (6.1) |  |
| CEE & medroxyprogesterone acetate<sup>1</sup> |  | 10 (3.2) |  |
| CEE & norethisterone<sup>1</sup> |  | 2 (0.6) |  |
| estradiol valerate & norethisterone<sup>1</sup> |  | 8 (2.5) |  |
| estradiol valerate & levonorgestrel<sup>2</sup> |  | 3 (1.0) |  |
| estradiol valerate & medroxyprogesterone acetate<sup>1</sup> |  | 3 (1.0) |  |
| estradiol & norethisterone acetate<sup>1</sup> |  | 7 (2.2) |  |
| estradiol & norethisterone<sup>1</sup> |  | 3 (1.0) |  |
| estradiol & progesterone<sup>o1</sup> |  | 2 (0.6) |  |
| tibolone |  | 13 (4.1) |  |
| mixed |  | 163 (51.9) |  |

\*Mean ± Standard Deviation. <sup>o</sup>Bioidentical form (no circle indicates synthetic form); <sup>1-3</sup> = progestin generations; <sup>l</sup>incl. subcutaneous and intravenous injections. Abbreviations: N, sample size; GCSE, General Certificate of Secondary Education; CSE, Certificate of Secondary Education; NVQ, National Vocational Qualification; BMI, body mass index; APOE, apolipoprotein; CEE, conjugated equine estrogen. Significant group differences are highlighted in bold.

### MRI data acquisition and processing

A detailed overview of the UK Biobank neuroimaging data acquisition and protocols has been published elsewhere ^46, 47^. Harmonized analysis pipelines were used to process raw T1-weighted MRI data for all participants (N = 20,540), including automated surface-based morphometry and subcortical segmentation (FreeSurfer, v5.3). To remove poor-quality data likely due to motion, participants with Euler numbers ≥ 4 standard deviations (SD) ^48^ below the mean were excluded (n = 180), yielding a total of 20,360 participants with T1-weighted MRI data (see *Sample characteristics* for final sample size).

In addition to the classic set of subcortical and cortical summary statistics from FreeSurfer ^49^, we utilized a fine-grained cortical parcellation scheme ^50^ to extract cortical thickness, area, and volume for 180 regions of interest per hemisphere. This yielded a total set of 1,118 structural brain imaging features (360/360/360/38 for cortical thickness/area/volume, as well as cerebellar/subcortical and cortical summary statistics, respectively), that were used to predict global GM brain age. The T1-weighted MRI data was residualized with respect to scanning site and intracranial volume using linear models ^51^. To probe hippocampal-specific effects separately ^52^, we used the extracted measures of left and right hippocampus volume as additional outcome measures in subsequent analyses.

Diffusion-weighted MRI data were processed using an optimized diffusion pipeline ^53–55^. Metrics from four diffusion models were utilized to predict global WM brain age (see details **supplementary Note 1**). In total, 98 diffusion features were included (global mean values + tract values for each diffusion metric). The diffusion-weighted data passed TBSS post-processing quality control using the YTTRIUM algorithm ^54^ and were residualized with respect to scanning sites using linear models.

Total volume of WMH was derived from T1-weighted and T2-weighted images using BIANCA (FSL, v6.0); a fully automated, supervised method for WMH detection ^56^. Preprocessed volumes per participant were exported from the UK Biobank, Field ID: 25781, and log-transformed due to a left-skewed distribution. A total of 19,538 females had data to compute WMH.

### Brain-age prediction

In line with our previous studies ^55, 57^, tissue-specific age prediction models in females only were run using *XGBoost* regression, which is based on a decision-tree ensemble algorithm (https://github.com/dmlc/xgboos). Hyper-parameters were tuned in nested cross-validations using 5 inner folds for randomized search, and 10 outer folds for model validation. Predicted age estimates were derived using the Scikit-learn library (https://scikit-learn.org), and brain age gap (BAG) values were calculated for each model (predicted – chronological age) to provide estimates of global GM BAG based on T1-weighted data, and global WM BAG based on diffusion-weighted data. The age prediction models were run without a subsample with ICD10 diagnosis known to impact the brain (n = 1,739, for details see **supplementary Note 2**), and then applied to the respective group of participants with diagnoses to obtain brain age estimates for the whole sample. This approach was selected to base the prediction models on normative age trajectories, while also including a more representative total sample (females both with and without diagnoses) in the subsequent analyses.

### MHT-related variables

For the whole sample, general MHT data included user status (never-user, current-user, or past-user), age first started using MHT, age last used MHT, and duration of MHT use (age last used – age first used). In current MHT users, age at last use was set to their age at the imaging assessment to calculate duration of use.

For a sub-sample, prescription MHT data was extracted from primary care records (general practice, UK Biobank Field ID: 42039) using freely available <u>code</u> ^58^. We extracted formulation (i.e., estrogens only, estrogens + progestins), route of administration (i.e., oral, transdermal, vaginal, injection), and daily drug dosage (mg) from the trade names/active ingredients indicated by the treating general practitioner and duration of use (weeks) was calculated based on prescription dates. Trade names were verified using the UK Product compendium (https://www.medicines.org.uk/emc). MHT formulations were further separated into bioidentical & synthetic, and progestins were stratified by generation (i.e., 1^st^ to 3^rd^ generation), based on their chemical structures, receptor binding properties, and clinical characteristics ^59^. Although all hormones in MHTs are chemically synthesized, bioidentical hormones are structurally identical to the hormones naturally produced in the human body, whereas synthetic hormones are not. It is debated whether bioidentical and synthetic hormones might have different risk profiles ^60^.

Drug duplicates, meaning the same drug issued on the same date per participant, were excluded. Individuals who switched between regimes were labeled as mixed (see **Table 3**). Individuals who took MHT after the imaging assessment, based on MHT prescription dates, were excluded (n = 6). In total, 538 participants had both detailed MHT and imaging data, and complete data on key demographic variables such as age, education, and menopausal status (see **Table 3**). Females who had missing data, or had responded “do not know”, “prefer not to answer”, “none of the above” or similar for any of the relevant variables, were excluded for the respective analyses. If possible and appropriate, missing data at the imaging time point was replaced with valid data from the baseline assessment. This was the case for the following variables: MHT user status, history of hysterectomy and/or bilateral oophorectomy, and menopausal status. For details on MHT-related variables in the UK Biobank see supplementary **Table S1**.

### APOE*ε* genotyping

For genotyping, we used the extensive quality control UK Biobank version 3 imputed data ^61^. The APOE ε genotype was approximated based on the two APOE ε single-nucleotide polymorphisms – rs7412 and rs429358 ^62^. APOE e4 status was labeled *carrier* for ε3/ε4 and ε4/ε4 combinations, and *non-carrier* for ε2/ε2, ε2/ε3 and ε3/ε3 combinations. Due to its ambiguity with ε1/ε3, the homozygous ε2/ε4 allele combination was removed (n = 484) ^63^ (https://www.snpedia.com/index.php/APOE). Further information on the genotyping process is available in the <u>UK Biobank documentation</u>.

### Statistical analyses

The statistical analyses were run using R, v4.2.2. P-values were corrected for multiple comparisons using false discovery rate (FDR) ^64^ correction across all brain measures for all sets of analyses per model (1-3). The sets of FDR corrections are reflected in the corresponding results tables. All variables were standardized prior to performing the regression analyses (subtracting the mean and dividing by the standard deviation). All statistical main and sensitivity analyses by sample and research questions are summarized in **Figure 1**.

**Figure 1.**
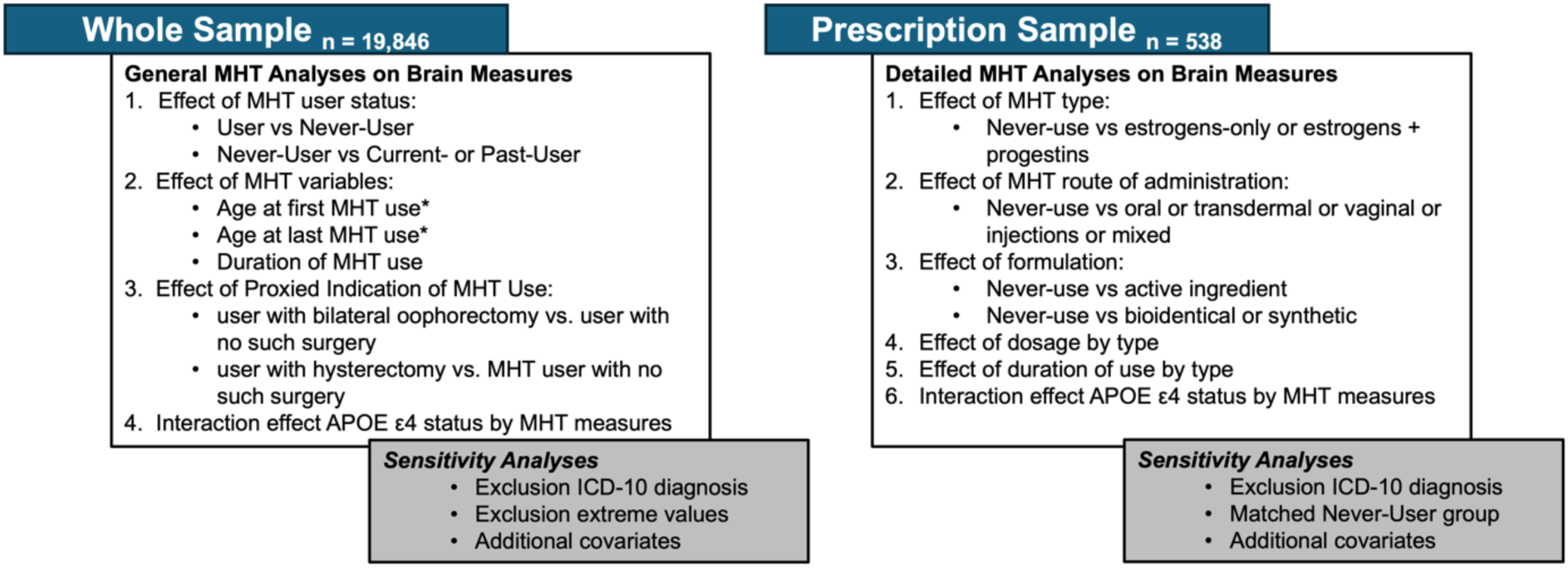
Overview of statistical main and sensitivity analyses by sample and research question.

### a. Associations between MHT variables and brain measures in the whole sample

First, we tested for associations between MHT user status (never-user/user) and GM and WM BAG, left and right hippocampus volume, and WMH volume. We further tested whether current and past MHT use, relative to never-use, was associated with the brain measures.

Second, among all MHT users, we assessed associations between age at first use as well as duration of use and brain measures. Among past MHT users, we tested for associations between age at last use and brain measures. In postmenopausal MHT users, we also tested whether age at first and last use in relation to age at menopause (i.e., age started MHT – age at menopause; age last use MHT – age at menopause, respectively) was associated with the brain measures. The following regression models (*lm* function) were fitted for these analyses, with DV representing each MRI measure (i.e., GM BAG, WM BAG, left and right hippocampus volumes, WMH volume) as dependent variable and IV representing each MHT variable (i.e., MHT user status, age at first use, age at last use, duration of use), as independent variable:

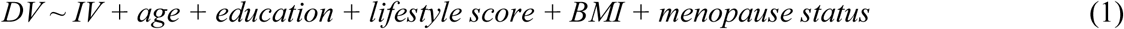

The chosen covariates are known to influence MHT use and brain structure ^65–68^. The lifestyle score was calculated using a published formula ^69^, and included data on sleep, physical activity, nutrition, smoking, and alcohol consumption (see supplementary **Note 3, Table S2**). For these analyses, participants with a history of hysterectomy and/or bilateral oophorectomy were excluded (n = 3,903), as they might have an increased risk for neurological decline ^70^. Note that Model 1 was not adjusted for menopause status in the analyses only including postmenopausal MHT users.

To test for differences in brain measures between MHT users with a history of hysterectomy without bilateral oophorectomy or bilateral oophorectomy (+/-hysterectomy, proxy of surgical menopause) relative to MHT users without such surgeries, we ran additional regression models within the sample of MHT users using the same model as specified above (model 1).

### b. Associations between MHT variables and brain measures in a subsample with prescription MHT data

First, we tested whether MHT formulation (i.e., estrogens-only, estrogens + progestins, none) and route of administration (i.e., oral, transdermal, vaginal, injections, mixed, none) was associated with brain measures. Never-users (dummy-coded as “none”) served as a reference group. The following regression model was fitted for these analyses, with DV representing each MRI measure and IV representing MHT formulation or route of administration:

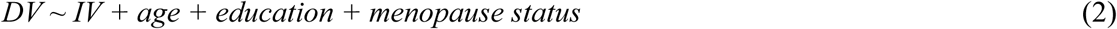

For these analyses, we only included age, education, and menopause status as covariates to retain the largest possible sample size (n = 538; see ‘Sensitivity Analyses’ for follow-up analyses including history of hysterectomy and/or bilateral oophorectomy as an additional covariate).

In estrogens-only users, we tested whether different active ingredients (i.e., estradiol valerate, estradiol hemihydrate, CEE, estradiol, mixed, none) and bioidentical or synthetic estrogens forms were associated with brain measures, relative to never-users as dummy-coded reference group. We further assessed whether duration of estrogens use (weeks), and estrogens drug dosage was associated with brain measures among estrogens-only users. We used the same model as specified above (model 2).

In estrogens + progestin users, we also explored whether different active ingredients were associated with brain measures, relative to never-users (see model 2). For this analysis, we only included formulations with N >= 10 (see **Table 3**). We further tested whether bioidentical or synthetic MHT or a mix of both, and progestin generations (i.e., 1^st^ or 2^nd^) were associated with brain measures, relative to never-users (see model 2). Third progestin generation users (n = 1) were excluded. Among estrogens + progestin users, we further assessed whether duration of estrogens/progestin use (weeks) and drug dosage of estrogens/progestin was differently associated with brain measures. For these models, the estrogens/progestin measures (i.e., duration of use or drug dosage) were included in the same model. The covariates specified in model 2 were included.

### c. Associations between MHT variables and brain measures by APOE ε4 status

First, we tested for the main effects of APOE ε4 status (i.e., carrier vs non-carrier) and APOE ε4 dose (i.e., non-carrier, ε3/ε4, ε4/ε4) on brain measures, in separate models:

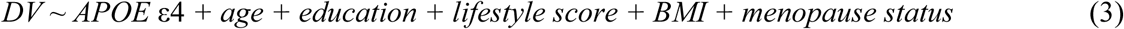

Similar to model 1, participants with a history of hysterectomy and/or bilateral oophorectomy were excluded for these analyses. Non-carrier served as a reference group.

Second, we re-ran models 1-2 including an APOE ε4 status × MHT measure interaction term to assess the effects of APOE ε4 status on the associations between MHT variables and brain measures. Main effects for the interaction terms were automatically included in the model.

## Sensitivity Analyses

To test whether the results were influenced by the inclusion of participants with ICD-10 diagnosis or by non-linear effects of age, the main analyses (models 1-2) were re-run excluding the sub-sample with diagnosed brain disorders (see supplementary **Note 2**) or adding age^2^ as additional covariate, respectively. In addition, we re-ran the analyses for MHT formulation and route of administration (Model 2) adjusting for a history of hysterectomy and/or bilateral oophorectomy in addition to age, BMI, and lifestyle score (n = 460). Since estrogens only MHTs are commonly prescribed after hysterectomy ± bilateral oophorectomy, we included surgical history as a covariate instead of excluding participants with a surgical history.

To adjust for the potential influence of extreme values on our results, we assessed each continuous MHT variable (i.e., age at first use, age at last use, age at menopause) for extreme values using a data-driven approach and excluded the corresponding participants before re-running the respective main analyses (model 1). To identify extreme values, we applied the median absolute deviation (MAD) method (R package *Routliers*), using default settings (i.e., a MAD threshold of ± 3).

For relevant analyses, the subsample with prescription MHT data was compared to all available never-users, to allow for a large and representative control group. However, to assess whether the results were sensitive to the control group selection, we matched the prescription MHT data sample (n = 538) to an equally sized subsample of never-users (n = 538), using genetic matching without replacement (*matchit* R package). Genetic matching is a form of nearest neighbor matching where distances are computed as the generalized Mahalanobis distance. The generalization of the Mahalanobis distance is achieved with a scaling factor for each covariate that represents the importance of that covariate to the distance. The groups were matched based on the covariates used in model 2, namely age, education, and menopause status. In the matched sample, for analyses comparing users relative to never-users, model 2 was rerun adjusting only for age.

## Results

### Sample characteristics

Sample demographics including lifestyle score, stratified by MHT user group, surgical history among MHT users, and estrogen only MHT or combined MHT use, are summarized in Table 1, 2 and 3, respectively. MHT user group differences for each lifestyle factor contained in the lifestyle score are shown in Table S2.

### Brain age prediction

The age prediction accuracy largely corresponded to our previous UK Biobank studies in overlapping samples ^21, 57^, as shown in **Table S3**. **Figure 2** shows the correlations between GM BAG, WM BAG, left and right hippocampus volume, and WMH volume.

**Figure 2.**
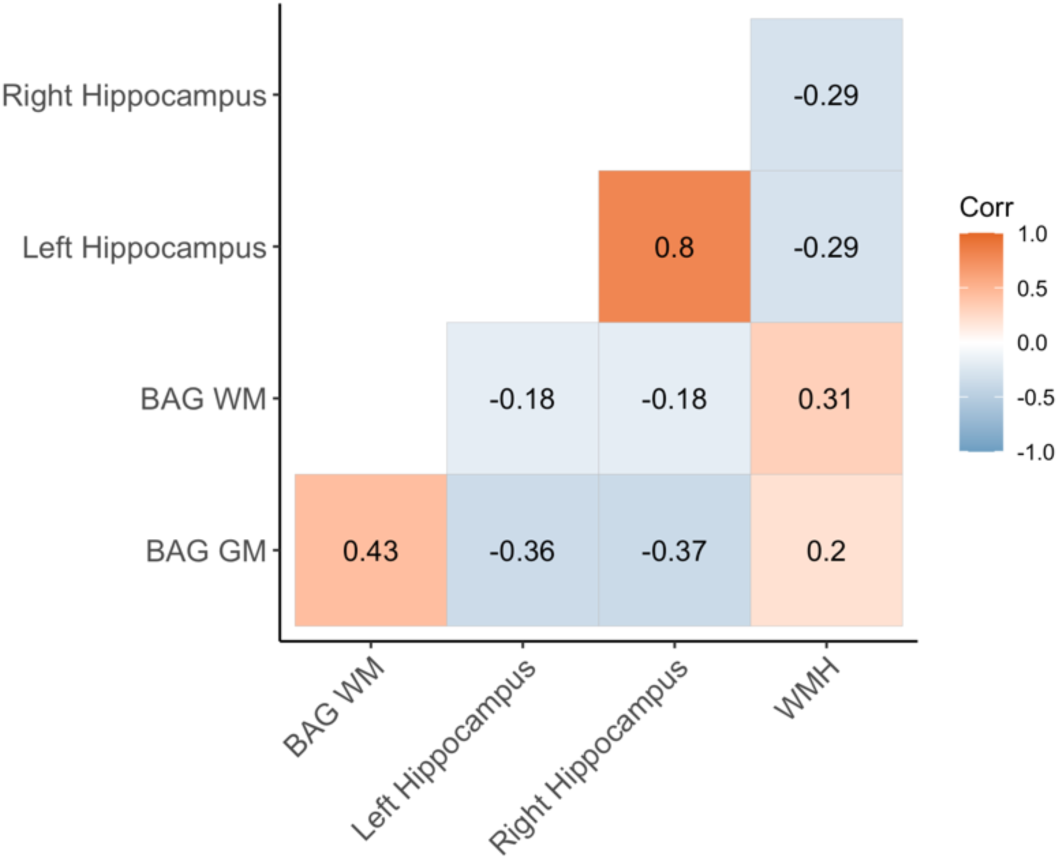
Correlations (Pearson’s r) between white matter (WM) and grey matter (GM) brain age gap (BAG) as well as left and right hippocampal volumes and total white matter hyperintensity (WMH) volume. BAG measures are adjusted for age^63^. WM BAG, GM BAG and hippocampal volumes were available for 20,360 individuals, and 19,538 had data on WMH volume.

### Associations between MHT variables and brain measures in the whole sample

In the whole sample, MHT users showed higher GM BAG (i.e., older brain age relative to chronological age; β=0.034, p=6.74e-05, p_FDR_ = 0.001) and lower left hippocampal volume (β=-0.02, p=0.006, p_FDR_=0.02) compared to never-users (**Table S4**). As shown in **Figure 3**, stratifying the sample into never-, past-, and current-users showed statistically significant higher GM and WM BAG and lower left and right hippocampal volumes in current users compared to never-users, but no significant difference in past-users relative to never-users. For WMH volume, there were no significant differences between current and past MHT users relative to never-users, respectively (**Table S4, Figure 3**).

**Figure 3.**
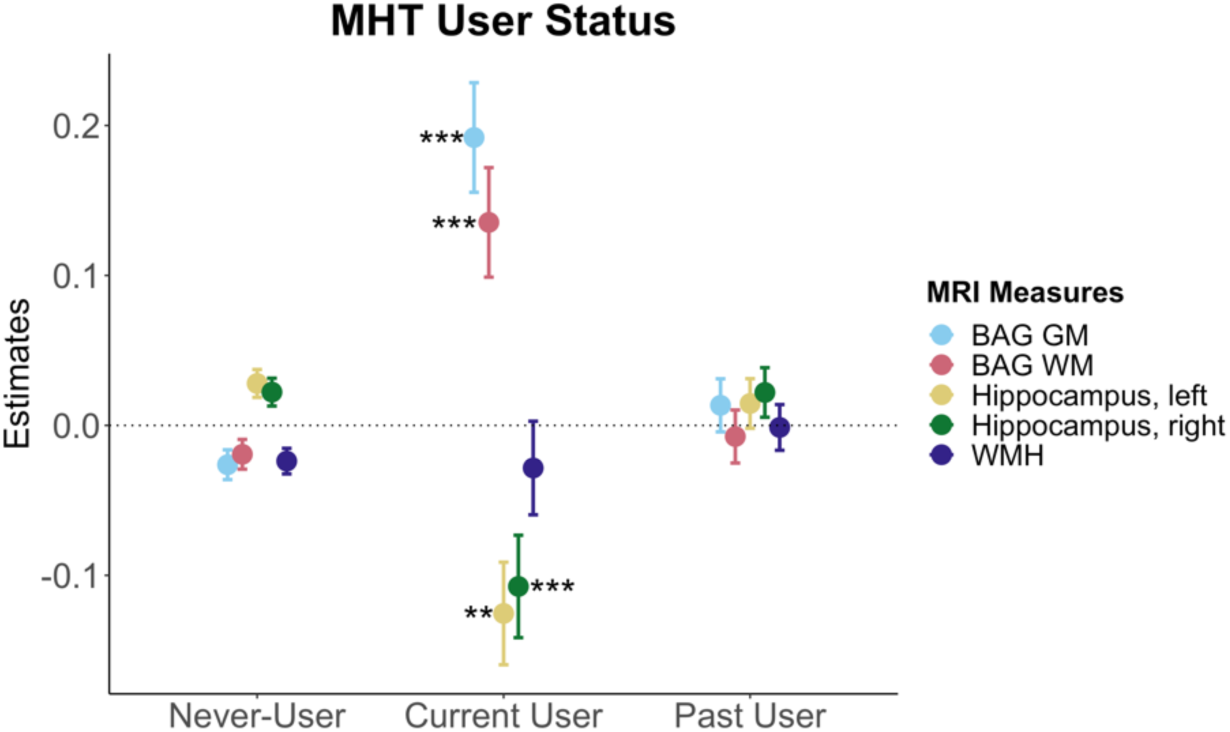
Associations between brain MRI measures and MHT user status. Point plot of estimated marginal means with standard errors from separate regression models with brain measure as dependent variable and MHT user status as independent (categorical) variable, with never-users as a reference group. Brain measures include white matter (WM) and grey matter (GM) brain age gap (BAG) as well as left and right hippocampal volumes and total white matter hyperintensity (WMH) volume. The models were adjusted for age, education, body mass index, lifestyle score, and menopausal status. All variables were standardized prior to performing the regression analysis (subtracting the mean and dividing by the standard deviation). Stars indicate significant associations. Significance codes: 0 ‘***’ 0.001 ‘**’ 0.01 ‘*’.

Among MHT users, we found no relationships between age at MHT initiation, alone and in relation to age at menopause, and the MRI variables (**Table S4)**. However, in past MHT users, older age at last use was associated with higher GM BAG. In postmenopausal past MHT users, older age at last use after age at menopause was associated with higher GM and WM BAG, higher WMH volume, and lower left and right hippocampal volumes. Similarly, longer duration of MHT use was associated with higher GM BAG and WM BAG, as well as lower left and right hippocampal volumes (**Table S4)**.

MHT use with a history of hysterectomy ± bilateral oophorectomy was associated with *lower* GM BAG (i.e., younger brain age relative to chronological age) relative to MHT users without such history. In addition, hysterectomy without oophorectomy was associated with higher left and right hippocampus volumes relative to MHT users without surgical history (**Figure 4**).

**Figure 4.**
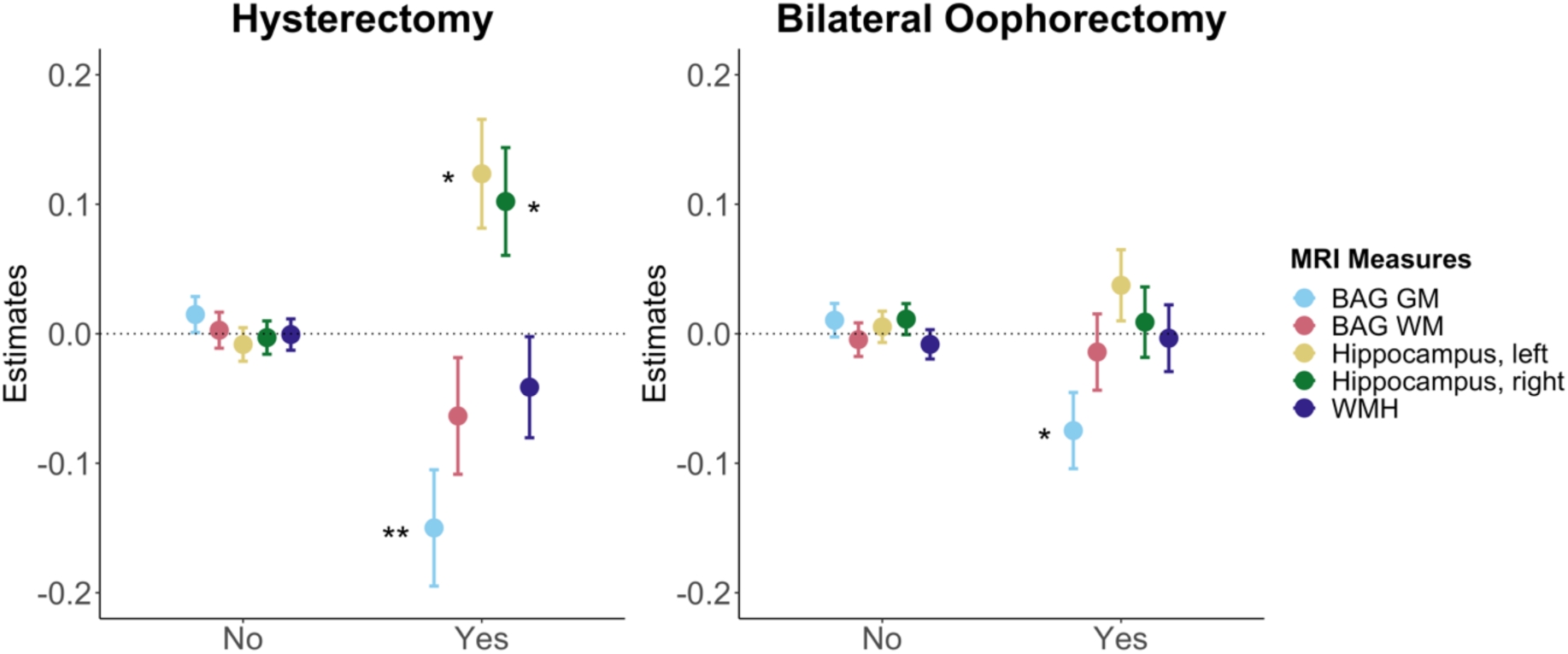
Associations between brain MRI measures and history of hysterectomy and/or bilateral oophorectomy in MHT users. Point plot of estimated marginal means with standard errors from separate regression analysis with MRI measure as dependent variable and history of hysterectomy and/or bilateral oophorectomy as independent (categorical) variable. MHT users without such surgical history served as a reference group. Brain measures include white matter (WM) and grey matter (GM) brain age gap (BAG) as well as left and right hippocampal volumes and total white matter hyperintensity (WMH) volume. The models were adjusted for age, education, body mass index, lifestyle score, and menopausal status. All variables were standardized prior to performing the regression analysis (subtracting the mean and dividing by the standard deviation). Stars indicate significant associations. Significance codes: 0 ‘***’ 0.001 ‘**’ 0.01 ‘*’.

### Associations between MHT variables and brain measures in a subsample with prescription MHT data

We found no significant associations between MHT formulation, route of administration, MHT type (i.e., bioidentical vs synthetic), MHT active ingredients, progestin generation, dosage, duration of use, and brain measures after adjusting for multiple comparisons (**Table S5**).

Before adjusting for multiple comparisons, we observed higher WM BAG in estrogens + progestin users relative to never-users (β=0.127, p=0.026, p_FDR_= 0.832). Users of CEE + MPA (β=0.634, p=0.045, p_FDR_= 0.994) as well as mixed active ingredients (β=0.179, p=0.023, p_FDR_= 0.832) showed higher WM BAG compared to never-users, and oral users showed higher WMH volume (β=0.134, p=0.020, p_FDR_=0.832). We further observed an association between longer duration of estrogens use and lower WMH volume in estrogens-only users relative to never-users (β=-0.146, p=0.028, p_FDR_=0.832). In estrogens + progestin users, longer duration of estrogens use was trend-level associated with lower left and right hippocampal volumes (left: β=-0.149, p=0.031, p_FDR_=0.832; right: β=-0.165, p=0.015, p_FDR_=0.832).

### Associations between MHT variables and brain measures by APOE ***ε***4 status

We found significantly lower right hippocampal volumes in carriers of two APOE ε4 alleles compared to non-carriers **(Figure 5).** We also observed higher GM BAG and lower left hippocampus volumes in APOE ε4-carriers relative to non-carriers (**Table S6**), but these effects were not significant after adjusting for multiple comparisons.

**Figure 5.**
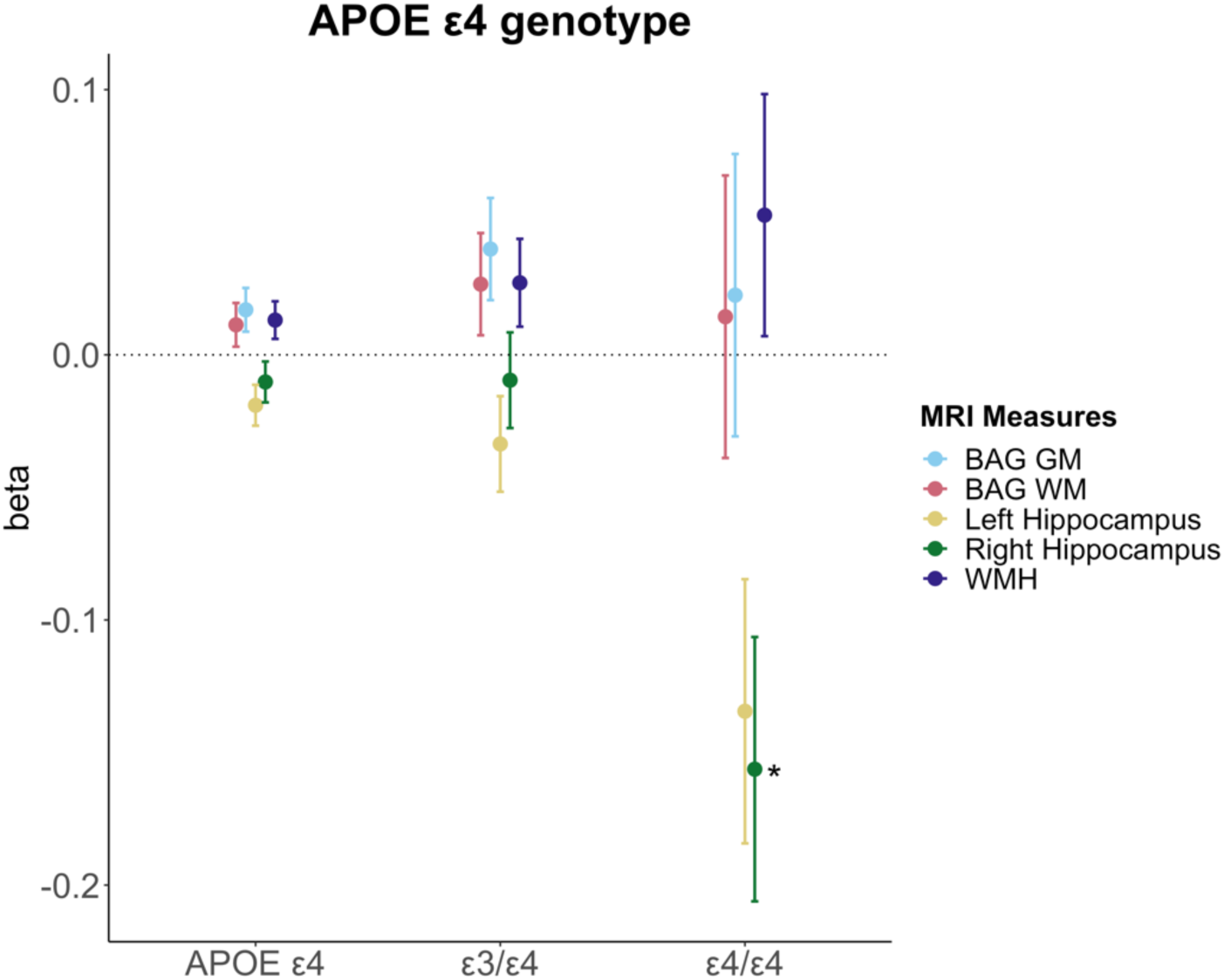
Associations between APOE ε4 genotype and brain MRI measures. Point plot of standardized beta-values with standard errors from separate multiple regression analysis with MRI measure as dependent variable and APOE ε4 genotype as independent variable. Non-carrier served as a reference group (n = 10,787). APOE ε4 (all, n = 3935) represents the ε3/ε4 (n = 3572) and ε4/ε4 carriers (n = 363) grouped together. For WMH, the sample sizes were as follows: non-carrier (n = 10,377), APOE ε4 (all, n = 3410), ε3/ε4 (n = 3410) and ε4/ε4 carriers (n = 349). Brain measures include white matter (WM) and grey matter (GM) brain age gap (BAG) as well as left and right hippocampal volumes and total white matter hyperintensity (WMH) volume. The models were adjusted for age, education, body mass index, lifestyle score, and menopausal status. All variables were standardized prior to performing the regression analysis (subtracting the mean and dividing by the standard deviation). Stars indicate significant associations. Significance codes: 0 ‘***’ 0.001 ‘**’ 0.01 ‘*’.

We found no significant interactions between APOE ε4 status and MHT variables on brain measures after adjusting for multiple comparisons, both for the analyses including the whole sample (**Table S7**) and for the MHT prescription sample (**Table S8**).

Before adjusting for multiple comparisons, we observed a trend-level interaction between APOE ε4 status and MHT use in participants with a hysterectomy without oophorectomy on WMH volume (β=-0.025, p=0.046, p_FDR_=0.927) in the whole sample. In the prescription sample, we observed several interactions between APOE ε4 status and MHT variables on brain measures before FDR correction. For example, mixed estrogens only use showed an interaction with APOE ε4 status on right hippocampus volume (β=-0.491, p=0.032, p_FDR_=0.511), indicative of smaller volumes in APOE ε4 carriers relative to non-carriers. We also observed an interaction between APOE ε4 status and synthetic estrogens + progestin use on GM BAG (β=-0.512, p=0.020, p_FDR_=0.449), left hippocampus volume (β=0.502, p=0.016, p_FDR_=0.449), and WMH volume (β=-0.412, p=0.029, p_FDR_=0.449), suggesting lower GM BAG, larger left hippocampus volume, and lower WMH volume in APOE ε4 carriers compared with non-carriers. Please see supplementary **Note S4** for the results of all models including APOE ε4 in the prescription sample.

## Sensitivity Analyses

The results were consistent after removing participants with ICD-10 diagnoses known to impact the brain (see **Table S9** for model 1 analyses and **Table S10** for model 2 analyses), after additionally adjusting for age^2^ (see Table S11), and after removing extreme values (see **Table S12** for model 1 analyses). Detected extreme values are highlighted in **Table S13**. Similarly, additionally adjusting for BMI, lifestyle score, and history of hysterectomy and/or bilateral oophorectomy (model 2, **Table S14**) or matching never-users to MHT users did not alter the results for the prescription dataset (model 2, **Table S15**).

## Discussion

This study assessed detailed MHT data, APOE ε4 genotype, and brain characteristics in a large, population-based sample of females in the UK. The results showed significantly higher GM and WM BAG (older brain age relative to chronological age) as well as smaller left and right hippocampus volumes in current MHT users, but not in past users, compared to never-users. Among MHT users, we found no significant associations between age at MHT initiation, alone and in relation to age at menopause, and brain measures. However, older age at last use after age at menopause was associated with higher GM and WM BAG, higher WMH volume, and lower left and right hippocampal volumes. Similar associations were found for longer duration of MHT use. MHT users with a history of hysterectomy ± bilateral oophorectomy showed *lower* GM BAG relative to MHT users without such history. In the sub-sample with prescription data, we found no significant associations between detailed MHT variables, such as MHT formulation, route of administration, type, active ingredients, and dosage, and brain measures, after FDR correction. Lastly, although we found lower right hippocampus volumes in carriers of two APOE ε4 alleles relative to non-carriers, we found no interactions with MHT variables after FDR correction.

Current MHT users showed higher GM and WM BAG as well as lower left and right hippocampus volumes relative to never-users. The effects were robust but relatively modest in magnitude, with the largest effect size indicating a group difference of 0.77 years (∼9 months) for GM BAG (standardized β=0.218, **Table S4**). However, we found no significant differences in brain measures in past MHT users relative to never-users. Current MHT users were significantly younger than past- and never-users, and around 67 % were menopausal relative to over 80% in the past- and never-user groups. The unequal distribution of age and menopausal status across groups may have influenced the observed findings. For instance, a larger proportion of the current users might be in the perimenopausal phase, which is often associated with debilitating neurological and vasomotor symptoms ^1^. MHT is commonly prescribed to minimize such symptoms. Although MHT initiation during perimenopause has been associated with improved memory and hippocampal function, as well as lower AD risk later in life ^15^, the need for MHT might in itself be an indicator of neurological changes ^71^; here potentially reflected in higher BAG and lower hippocampal volumes. After the transition to menopause, symptoms might subside and some perimenopausal brain changes might revert or stabilize in the postmenopausal phase ^5^. Although the UK Biobank lacks detailed information on menopausal symptoms and perimenopausal staging, our results might be capturing subtle disturbances during perimenopause that later stabilize. This could explain why the largely postmenopausal groups of past MHT users and never-users present with lower GM and WM BAG than the current user group. Considering the critical window hypothesis emphasizing perimenopause as a key phase for MHT action ^29, 43^, future longitudinal studies are crucial to clarify the interplay between neurological changes and MHT use across the menopause transition.

Besides age-related differences, the current user group also showed a significantly unhealthier lifestyle, and higher rates of bilateral oophorectomy compared to the never-user group and the past-user group, respectively. These findings are contrary to the healthy user bias hypothesis (i.e., equating MHT use with healthy user status ^72^), and might indicate that the current user group could be exposed to neurological changes prior to MHT use. According to the healthy cell bias hypothesis of estrogen action ^73^, MHT use might be detrimental for brain health when initiated after cells are exposed to neurological degeneration. Although MHT use might have exacerbated adverse brain changes in the unhealthier group of current users, higher BAG was also linked to longer duration of MHT use and older age at last use post menopause. Although the effect sizes were modest (**Table S4**), these findings might reflect subtle yet unfavorable effects of MHT on brain health in our sample, particularly when used continuously after natural menopause with uterus and ovaries still intact (i.e., all females with hysterectomy ± bilateral oophorectomy were excluded from this analysis).

To the contrary, MHT users with a history of hysterectomy ± bilateral oophorectomy showed *lower* GM BAG relative to MHT users without such history, and we found larger hippocampal volumes in MHT users with a hysterectomy without a bilateral oophorectomy. Previous work associates these surgical interventions with accelerated cognitive decline ^74^, increased risk of dementia ^70^, morphological changes in regions of the medial temporal lobe ^75^, and accumulation of Alzheimer’s disease pathology ^76^. However, some of these studies relied on comparing surgery to no surgery ^70, 74^ without taking MHT use into account ^77^, or investigated the effect of surgery with MHT relative to surgery without MHT. For instance, a 2023 study highlighted that estrogen-only use in females with hysterectomy was associated with increased dementia rates relative to females with hysterectomy who never used MHT ^78^. In the current study, we specifically compared the effect of MHT use on brain measures among females with and without surgical history and found lower GM BAG in the surgical MHT user group. These findings might be explained by differences in indication of MHT use for females with and without surgery, MHT formulation, and age at surgery. Without surgery, MHT use (i.e., often combined MHT formulation) is prescribed in females with intact uterus and ovaries to minimize menopausal symptoms that are largely neurological in nature, such as vasomotor symptoms as well as mood, cognitive, and sleep disturbances ^1^. After hysterectomy, estrogen only MHT might be prescribed but is not strictly needed as ovaries are still intact. Whereas after bilateral oophorectomy (= surgical menopause), combined or estrogen only MHT is indicated to compensate for the acute and chronic deficiency of hormones normally produced by the ovaries. These distinctions might entail important differences in risk and side-effect profiles of estrogen only or combined MHT in females with and without endogenous sex hormone production. It is also possible that the timing between MHT use and surgery is more tightly controlled and therefore more beneficial for brain aging ^43^. For instance, studies suggest that MHT may mitigate the potential long-term adverse effects of bilateral oophorectomy before natural menopause on bone mineral density as well as cardiovascular, cognitive and mental health^79–81^. In addition, a 2024 UK Biobank study found that ever used MHT was associated with decreased odds of Alzheimer’s disease in women with bilateral oophorectomy^82^.

However, our study also showed group differences in several demographic and MHT-related variables which might influence the results. For instance, the hysterectomy MHT user group was significantly older than the non-surgery and bilateral oophorectomy group, started MHT at a younger age than the non-surgery group, and had surgery at a younger age than the bilateral oophorectomy group. More research is needed to disentangle this complex interplay of MHT effects in females with and without intact uteri and ovaries.

No significant associations were observed between brain measures and MHT regimes based on prescription data. However, given the relatively small sample size and the large number of comparisons, it is possible that we were unable to detect subtle effects of factors such as MHT formulation, route of administration, type, active ingredients, and dosage. Before FDR correction, we found higher WM BAG in estrogens + progestin users, in CEE + MPA users, and in mixed active ingredients users relative to never-users. In addition, WMH volume was higher among oral MHT users and lower with longer duration of estrogens use in estrogens-only users relative to never-users. These uncorrected results are partly in line with previous findings. For instance, the 2003 Women’s Health Initiative Memory Study reported that prolonged oral use of both CEE alone ^24^, or combined with MPA ^26^, increased the risk of dementia and cognitive decline among females aged 65 years and older. Contrarily, prolonged use of estrogen only MHT has been linked to reduced white matter loss in aging (n = 10) ^19^. Similar to our findings, Ha and colleagues did not find an association between gray matter volumes and estrogen use ^19^. Although uncorrected results must be interpreted with caution, our findings might indicate an unfavorable effect of mixed active ingredient use, including CEE ± MPA and oral administration, on white matter brain aging. This effect might be due to the higher estrone concentrations associated with such oral estrone-based MHT types ^83–85^. In addition, over 50% of combined MHT user had mixed MHT use which suggests that it might be beneficial to examine the reasons for switching of medication. In sum, these findings highlight the need for personalized MHT regimes, and longitudinal RCTs are needed to establish how different MHT regimes, and their respective hormonal profiles are causally linked to brain aging.

In the current study, we found significantly smaller right hippocampus volumes in carriers of two APOE ε4 alleles relative to non-carriers, which is in line with previous work linking the APOE ε4 genotype to greater rates of hippocampal atrophy in non-demented and Alzheimer’s disease samples ^86, 87^. However, after FDR correction, we did not find any interactions between APOE ε4 carrier status and MHT variables in relation to the brain measures. This finding was unexpected, as previous work has highlighted the APOE ε4 genotype as a crucial determinant of MHT effects on the female brain ^88^. For instance, one study associated MHT use with improved memory performance and larger entorhinal and amygdala volumes in female ε4 carriers versus non-carriers ^89^. A 2023 prospective longitudinal study showed that MHT was associated with smaller changes towards Alzheimer’s disease pathophysiology than non-therapy and that APOE ε4 carrier status was linked to an amplified treatment outcome ^90^. However, contrary results have also been reported. For instance, Yaffe and colleagues found lower risk of cognitive decline with estrogen only MHT use among female APOE ε4 non-carriers, but there was no such effect among carriers ^39^. To understand these discrepancies, more research on MHT by genotype interactions is needed.

The current work represents the most comprehensive study of detailed MHT data, APOE ε4 genotype, and several brain measures in a large population-based cohort to date. Overall, our findings do not unequivocally support general neuroprotective effects of MHT, nor do they indicate severe adverse effects of MHT use on the female brain. The results suggest subtle yet complex relationships between MHT’s and brain health, highlighting the necessity for a personalized approach to MHT use. Importantly, our analyses provide a broad view of population-based associations and are not designed to guide individual-level decisions regarding the benefits versus risks of MHT use. Furthermore, several study limitations should be acknowledged. The presented data does not enable causal inference, and observational studies are subject to different sources of heterogeneity such as switching between MHT regimes (e.g., due to side effects or availability) and variable MHT formulation and dose. In addition, utilizing prescription registry data comes with its own set of challenges. In the UK, a national system for collecting or sharing primary care data is currently missing, and the availability, completeness, and level of detail in the data might vary between systems, suppliers and over time. The primary care data used in the current study was drawn from an interim release, including data on approximately 231,000 participants. Hence, our analyses conducted on these participants’ data and the observed results might not generalize to the entire cohort. Furthermore, although we extracted prescription MHT data, medications that were prescribed were not necessarily dispensed or used. Moving forward, research utilizing unified prescription registry data is needed to overcome some of these hurdles. In addition, previous studies highlight that UK Biobank participants are considered healthier than the general population based on several lifestyle and health-related factors ^91, 92^. This healthy volunteer bias increases with age, likely resulting in a disproportionate number of healthier older adults. Together with the imbalance in age distributions across groups, this might explain the less apparent brain aging in the older MHT user groups. We have previously highlighted that age is negatively associated with the number of APOE ε4 carriers in the UK Biobank ^21^, which is indicative of survivor bias. In addition to these inherent biases in aging cohorts, the ethnic background of the sample is homogeneous (> 96% white), further reducing the generalizability of the results. Lastly, although the UK Biobank has a wealth of female-specific variables, the acquired data relies on self-reports, which might not be reliable and data recording does not always align with best practice standards. For example, menopausal status in the UK Biobank is recorded based on whether the menstrual period has generally stopped, not whether it has been absent for at least 12 months, in line with the STRAW criteria ^93^.

In conclusion, our findings suggest that associations between MHT use and female brain health might vary depending on duration of use and past surgical history. Although the effect sizes were generally modest, future longitudinal studies and RCTs, particularly focused on the perimenopausal transition window, are warranted to fully understand how MHT use influences female brain health. Importantly, considering risks and benefits, decisions regarding MHT use should be made within the clinical context unique to each individual.

## Acknowledgments

We thank Dr. Melis Anatürk for sharing her prescription registry data extraction script, Brian F. O’Donnell for assistance with prescription data extraction, Dr. Dennis van der Meer for the extraction of the APOE e4 genotype, Prof. Tobias Kaufman, and Dr. Ivan I. Maximov for establishing the MRI preprocessing infrastructure used for brain age prediction, and Dr. Caitlin Taylor for her insights on female health at the beginning of the project.

This research has been conducted using the UKB under Application 27412. The analyses were performed on the Service for Sensitive Data (TSD) platform, owned by the University of Oslo, operated, and developed by the TSD service group at the University of Oslo IT-Department (USIT). Computations were also performed using resources provided by UNINETT Sigma2 - the National Infrastructure for High Performance Computing and Data Storage in Norway.

## Data Availability Statement

The data that support the findings of this study are available through the UK Biobank application procedure (https://www.ukbiobank.ac.uk/enable-your-research/register); scripts are available from the authors upon request.

