## Supplementary Material for "Menopausal hormone therapy and the female brain: leveraging neuroimaging and prescription registry data from the UK Biobank cohort"

**Note 1| White Matter Brain Age Estimation**

Metric from four diffusion models were utilized to predict white matter brain age, namely diffusion tensor imaging (DTI)[1], diffusion kurtosis imaging (DKI)[2], white matter tract integrity (WMTI)[3], and spherical mean technique (SMT)[4, 5]. The DTI metrics included mean diffusivity (MD), fractional anisotropy (FA), axial diffusivity (AD), and radial diffusivity (RD)[1]. The DKI metrics included mean kurtosis (MK), axial kurtosis (AK), and radial kurtosis (RK)[2]. WMTI metrics included axonal water fraction (AWF), extra-axonal axial diffusivity (axEAD), and extra-axonal radial diffusivity (radEAD)[3]. SMT metrics included intra-neurite volume fraction (INVF), extra-neurite mean diffusivity (exMD), and extra-neurite radial diffusivity (exRD)[4]. For each diffusion model metric, WM features were extracted based on John Hopkins University (JHU) atlases for white matter tracts (with 0 thresholding)[6], including mean values and regional measures for 12 tracts[7-9]: anterior thalamic radiation (ATR), corticospinal tract (CST) cingulate gyrus (CG), cingulum hippocampus (CING), forceps major (FMAJ), forceps minor (FMIN), inferior fronto-occipital fasciculus (IFOF), inferior longitudinal fasciculus (ILF) superior longitudinal fasciculus (SLF), uncinate fasciculus (UF), superior longitudinal fasciculus temporal (SLFT), and corpus callosum (CC).

**Note 2| ICD-10 Diagnosis**

We excluded participants with diagnosed brain disorders (n = 1,739) from the brain age prediction and in sensitivity analyses, established based on ICD-10 criteria, including field F ('Mental and behavioral disorders') with F00-F03 for Alzheimer’s disease and dementia and F06.7 (´Mild cognitive disorder`), field G (`Diseases of the nervous system´) with inflammatory and neurodegenerative diseases (except G55-59; ´Nerve, nerve root and plexus disorders´) and field I (‘Diseases of the circulatory system’) with I64 for stroke. An overview of the diagnoses is provided in the UK Biobank online resources (https://biobank.ndph.ox.ac.uk/showcase/field.cgi?id=41270), and the diagnostic criteria are listed in the ICD10 diagnostic manual (https://www.who.int/classifications/icd/icdonlineversions).

**Note 3| Lifestyle Score**

The lifestyle score was calculated based on sleep duration, time spent watching television, current and past smoking status, alcohol consumption frequency, physical activity level (number of days per week of moderate/vigorous activity for at least 10 minutes), intake of fruits and vegetables, and intake of oily fish, beef, lamb/mutton, pork and processed meat (for details see [10]). Each unhealthy lifestyle factor was scored with 1 point (e.g., smoking), and participants points were summed to generate an unweighted score (from 0-9): the higher the lifestyle score, the unhealthier the participant’s lifestyle.

A comparison of the lifestyle factors contained in the lifestyle score by MHT user status is presented in Table S2. In summary, we found that current MHT were more often smokers than never-users, had a higher alcohol intake than never- and past MHT users, reported the lowest fruit and vegetable intake relative to never-users and past MHT users, and stated lower moderate activity levels relative to past MHT users. Past MHT users reported higher alcohol intake than never-users, spend more time watching TV relative to never- and current-users, consumed more beef, pork, lamb/mutton, and processed meat than never-users, and reported lower vigorous activity levels relative to never-users. However, oily fish intake and fruit and vegetable intake was higher among past MHT users relative to never-and current-users. Self-reported sleep duration did not differ between MHT user groups.

**Note 4| Detailed results in the prescription sample not surviving correction for multiple comparison.**

In the prescription sample, we observed several interactions between APOE ε4 status and MHT variables on MRI measures before adjusting for multiple comparisons. Mixed bioidentical & synthetic estrogens + progestin use also showed an interaction with APOE ε4 genotype on right hippocampus volume (β=-0.292, p=0.007, $p_{FDR}$=0.449). Among synthetic estrogens + progestin users, CEE & norgestrel use showed an interaction with APOE ε4 genotype on GM BAG (β=-0.513, p=0.042, $p_{FDR}$=0.561) and WMH volumes (β=-0.499, p=0.022, $p_{FDR}$=0.449). Tibolone use (β=0.881, p=0.044, $p_{FDR}$=0.561) and estradiol hemihydrate & norethisterone acetate use (β=-0.320, p=0.022, $p_{FDR}$=0.449) both showed an interaction with APOE ε4 genotype on left and right hippocampus volume, respectively. First generation progestin use (β=-0.231, p=0.018, $p_{FDR}$=0.449) and mean progestin dosage (β=0.512, p=0.010, $p_{FDR}$=0.449) also interacted with APOE ε4 genotype on right hippocampus volume and WMH volume, respectively. Concerning mode of administration, injection showed an interaction with APOE ε4 genotype on left hippocampus volume (β= -0.703, p=0.022, $p_{FDR}$=0.449).

**Table S1| Assessment of menopausal hormone therapy (MHT)-related variables in the UK Biobank (UKB).**

| MHT variable | UKB Data-field | ACE touchscreen question | Coding |
|---|---|---|---|
| Menopause | 2724 | “Have you had your menopause (periods stopped)?” | 1 (“Yes”)<br>0 (“No”)<br>2 («Not sure – had a hysterectomy»)<br>3 (“Not sure – other reason “)<br>-3 («Prefer not to answer») |
| Age at Menopause | 3581 | “How old were you when your periods stopped?” | Age in years<br>-1 («Do not know»)<br>-3 («Prefer not to answer») |
| Bilateral Oophorectomy | 2834 | “Have you had BOTH ovaries removed?” | 1 (“Yes”)<br>0 (“No”)<br>-5 («Not sure»)<br>-3 («Prefer not to answer») |
| Hysterectomy | 3591 | “Have you had a hysterectomy (womb removed)?” | 1 (“Yes”)<br>0 (“No”)<br>-5 («Not sure»)<br>-3 («Prefer not to answer») |
| MHT use | 2814 | “Have you ever used hormone replacement therapy (HRT)?” | 1 (“Yes”)<br>0 (“No”)<br>-1 («Do not know»)<br>-3 («Prefer not to answer») |
| Age at last MHT use | 3546 | “How old were you when you last used HRT?” | Age in years<br>-1 («Do not know»)<br>-11 (“Still taking HRT”)<br>-3 («Prefer not to answer») |
| Age at first MHT use | 3536 | “How old were you when you first used HRT?” | Age in years<br>-1 («Do not know»)<br>-3 («Prefer not to answer») |

**Table S2| Lifestyle factors, constituting the lifestyle score, in menopausal hormone therapy (MHT) never-, current, and past- users in the whole sample.**

| | MHT User Status | | | p-value | | |
|---|---|---|---|---|---|---|
| | **Never** | **Current** | **Past** | **Never vs Current** | **Never vs Past** | **Current vs Past** |
| **N** | 12,012 | 1,153 | 6,681 | | | |
| **Age (years) *** | 61.6 ± 7.1 | 60.1 ± 6.8 | 67.5 ± 6.2 | **<0.001** | **<0.001** | **<0.001** |
| **Smoking Status, Yes, N(%)** | 313 (2.7) | 43 (3.8) | 187 (2.9) | **0.032** | 0.387 | 0.118 |
| **Alcohol Intake, N(%)** | | | | **<0.001** | **<0.001** | **0.002** |
| Daily or almost daily | 1428 (12.1) | 181 (16.0) | 1012 (15.7) | | | |
| Three or four times a week | 2957 (25.2) | 316 (28.0) | 1581 (24.5) | | | |
| Once or twice a week | 3271 (27.8) | 306 (27.1) | 1662 (25.7) | | | |
| One to three times a month | 1654 (14.1) | 141 (12.5) | 808 (12.5) | | | |
| Special occasions only | 1572 (13.4) | 124 (11.0) | 905 (14.0) | | | |
| Never | 873 (7.4) | 61 (5.4) | 495 (7.7) | | | |
| **Time spend watching TV (hours)*** | 2.1 ± 3.0 | 2.1 ± 3.1 | 2.8 ± 2.5 | 0.720 | **<0.001** | **<0.001** |
| **Sleep duration (hours)*** | 7.1 ± 1.0 | 7.1 ± 1.1 | 7.1 ± 1.1 | 0.960 | 0.432 | 0.750 |
| **Fruit & Vegetable Intake (gram)*** | 685.1 ± 362.7 | 660.1 ± 342.1 | 706.0 ± 350.2 | **0.026** | **<0.001** | **<0.001** |
| **Oily Fish Intake, N(%)** | | | | 0.202 | **<0.001** | **<0.001** |
| Never | 1156 (9.8) | 113 (10.0) | 444 (6.9) | | | |
| Less than once a week | 3435 (29.2) | 321 (28.4) | 1568 (24.3) | | | |
| Once a week | 4795 (40.8) | 431 (38.2) | 2823 (43.7) | | | |
| 2-4 times a week | 2261 (19.2) | 252 (22.3) | 1572 (24.3) | | | |
| 5-6 times a week | 82 (0.7) | 9 (0.8) | 43 (0.7) | | | |
| Once or more daily | 26 (0.2) | 3 (0.3) | 13 (0.2) | | | |
| **Beef Intake, N(%)** | | | | 0.165 | **<0.001** | 0.053 |
| Never | 1851 (15.7) | 166 (14.7) | 800 (12.4) | | | |
| Less than once a week | 5834 (49.6) | 529 (46.9) | 3323 (51.4) | | | |
| Once a week | 3095 (26.3) | 327 (29.0) | 1750 (27.1) | | | |
| 2-4 times a week | 966 (8.2) | 107 (9.5) | 582 (9.0) | | | |
| 5-6 times a week | 8 (0.1) | 0 (0.0) | 6 (0.1) | | | |
| Once or more daily | 1 (0.0) | 0 (0.0) | 2 (0.0) | | | |
| **Pork Intake, N(%)** | | | | 0.962 | **<0.001** | **0.041** |
| Never | 2708 (23.0) | 260 (23.0) | 1322 (20.5) | | | |
| Less than once a week | 7114 (60.5) | 691 (61.2) | 3871 (59.9) | | | |
| Once a week | 1703 (14.5) | 158 (14.0) | 1128 (17.5) | | | |
| 2-4 times a week | 223 (1.9) | 20 (1.8) | 137 (2.1) | | | |
| 5-6 times a week | 5 (0.0) | 0 (0.0) | 3 (0.0) | | | |
| Once or more daily | 2 (0.0) | 0 (0.0) | 2 (0.0) | | | |
| **Lamb/Mutton Intake, N(%)** | | | | **0.048** | **<0.001** | 0.815 |
| Never | 3030 (25.8) | 261 (23.1) | 1418 (21.9) | | | |
| Less than once a week | 7293 (62.0) | 699 (61.9) | 4043 (62.6) | | | |
| Once a week | 1314 (11.2) | 155 (13.7) | 926 (14.3) | | | |
| 2-4 times a week | 116 (1.0) | 14 (1.2) | 76 (1.2) | | | |
| Once or more daily | 2 (0.0) | 0 (0.0) | 0 (0.0) | | | |
| **Processed Meat Intake, N(%)** | | | | 0.938 | **<0.001** | 0.073 |
| Never | 1741 (14.8) | 171 (15.1) | 803 (12.4) | | | |
| Less than once a week | 4832 (41.1) | 465 (41.2) | 2815 (43.6) | | | |
| Once a week | 2987 (25.4) | 280 (24.8) | 1661 (25.7) | | | |
| 2-4 times a week | 2017 (17.2) | 198 (17.5) | 1086 (16.8) | | | |
| 5-6 times a week | 147 (1.3) | 11 (1.0) | 87 (1.3) | | | |
| Once or more daily | 31 (0.3) | 4 (0.4) | 11 (0.2) | | | |
| **Moderate Activity (days/week)*** | 4.0 ± 2.3 | 3.9 ± 2.3 | 4.0 ± 2.4 | 0.126 | 0.051 | **0.018** |
| **Vigorous Activity (days/week)*** | 1.8 ± 1.8 | 1.8 ± 1.8 | 1.7 ± 1.9 | 0.312 | **<0.001** | 0.371 |

* Mean ± Standard Deviation. Fruit & Vegetable Intake is a composite of intake of dried fruits, fresh fruits, salad, raw vegetables and cooked vegetables. Smoking status take past and current smoking into account. Abbreviations: N, sample size. Significant differences between groups based on t/$\chi 2$ tests are highlighted in bold.

**Table S3| Age prediction accuracy for the global grey and white matter models.**

| Model | $R^2$ | RMSE | MAE | *r* [95% CI] | *p-value* |
|---|---|---|---|---|---|
| GM | 0.53 ± 0.02 | 5.03 ± 0.09 | 4.02 ± 0.09 | 0.73 [0.73,0.74] | **< 0.001** |
| WM | 0.45 ± 0.03 | 5.40 ± 0.08 | 4.35 ± 0.08 | 0.67 [0.66, 0.68] | **< 0.001** |

Abbreviations: $R^2$ = average $R^2$, RMSE = root mean square error, MAE = mean absolute error, r = correlation between predicted and chronological age, CI = confidence interval.

**Table S4| Associations between menopausal hormone therapy (MHT)-related variables and brain measures in the whole sample.**

| MHT Variable | MRI Measure | beta | S.E. | t-value | p-value | pFDR-value |
|---|---|---|---|---|---|---|
| MHT Status | GM BAG | 0.034 | 0.008 | 3.986 | **6.74e-05** | **0.001** |
| | WM BAG | 0.019 | 0.008 | 2.212 | **0.027** | 0.064 |
| | Left Hippocampus | -0.021 | 0.008 | -2.726 | **0.006** | **0.020** |
| | Right Hippocampus | -0.012 | 0.008 | -1.479 | 0.139 | 0.224 |
| | WMH | 0.008 | 0.007 | 1.078 | 0.281 | 0.402 |
| Current MHT use | GM BAG | 0.218 | 0.038 | 5.782 | **7.54e-09** | **3.77e-07** |
| | WM BAG | 0.155 | 0.038 | 4.101 | **4.13e-05** | **4.13e-04** |
| | Left Hippocampus | -0.153 | 0.035 | -4.344 | **1.41e-05** | **2.11e-04** |
| | Right Hippocampus | -0.130 | 0.035 | -3.674 | **2.39e-04** | **0.001** |
| | WMH | -0.005 | 0.032 | -0.142 | 0.887 | 0.985 |
| Past MHT use | GM BAG | 0.040 | 0.021 | 1.916 | 0.055 | 0.111 |
| | WM BAG | 0.012 | 0.021 | 0.574 | 0.566 | 0.707 |
| | Left Hippocampus | -0.013 | 0.019 | -0.690 | 0.490 | 0.645 |
| | Right Hippocampus | 0.000 | 0.019 | -0.012 | 0.990 | 0.992 |
| | WMH | 0.022 | 0.018 | 1.263 | 0.207 | 0.313 |
| Age at first MHT use | GM BAG | 0.003 | 0.016 | 0.221 | 0.825 | 0.960 |
| | WM BAG | 0.000 | 0.015 | -0.010 | 0.992 | 0.992 |
| | Left Hippocampus | 0.010 | 0.015 | 0.669 | 0.503 | 0.645 |
| | Right Hippocampus | -0.001 | 0.015 | -0.067 | 0.946 | 0.986 |
| | WMH | -0.024 | 0.013 | -1.797 | 0.072 | 0.129 |
| Age at first MHT use relative to age at menopause | GM BAG | 0.028 | 0.017 | 1.682 | 0.093 | 0.160 |
| | WM BAG | 0.031 | 0.016 | 1.889 | 0.059 | 0.114 |
| | Left Hippocampus | 0.002 | 0.016 | 0.098 | 0.922 | 0.986 |
| | Right Hippocampus | -0.029 | 0.016 | -1.845 | 0.065 | 0.121 |
| | WMH | -0.007 | 0.014 | -0.494 | 0.622 | 0.758 |
| Age at last MHT use | GM BAG | 0.046 | 0.018 | 2.550 | **0.011** | **0.030** |
| | WM BAG | 0.039 | 0.018 | 2.165 | **0.030** | 0.069 |
| | Left Hippocampus | -0.036 | 0.018 | -2.036 | **0.042** | 0.091 |
| | Right Hippocampus | -0.027 | 0.017 | -1.565 | 0.118 | 0.196 |
| | WMH | 0.019 | 0.016 | 1.211 | 0.226 | 0.332 |
| Age at last MHT use relative to age at menopause | GM BAG | 0.062 | 0.018 | 3.413 | **0.001** | **0.003** |
| | WM BAG | 0.064 | 0.018 | 3.505 | **4.63e-04** | **0.002** |
| | Left Hippocampus | -0.055 | 0.018 | -3.100 | **0.002** | **0.008** |
| | Right Hippocampus | -0.056 | 0.018 | -3.205 | **0.001** | **0.006** |
| | WMH | 0.039 | 0.016 | 2.440 | **0.015** | **0.039** |
| Duration of MHT use | GM BAG | 0.072 | 0.016 | 4.454 | **8.67e-06** | **2.11e-04** |
| | WM BAG | 0.062 | 0.016 | 3.853 | **1.19e-04** | **0.001** |
| | Left Hippocampus | -0.065 | 0.015 | -4.307 | **1.69e-05** | **2.11e-04** |
| | Right Hippocampus | -0.044 | 0.015 | -2.915 | **0.004** | **0.012** |
| | WMH | 0.027 | 0.014 | 1.924 | 0.054 | 0.111 |
| Bilateral Oophorectomy | GM BAG | -0.033 | 0.013 | -2.594 | **0.009** | **0.028** |
| | WM BAG | -0.004 | 0.013 | -0.291 | 0.771 | 0.918 |
| | Left Hippocampus | 0.012 | 0.012 | 1.046 | 0.296 | 0.411 |
| | Right Hippocampus | -0.001 | 0.012 | -0.076 | 0.939 | 0.986 |
| | WMH | 0.002 | 0.011 | 0.164 | 0.869 | 0.985 |
| Hysterectomy | GM BAG | -0.047 | 0.013 | -3.497 | **4.74e-04** | **0.002** |
| | WM BAG | -0.019 | 0.014 | -1.401 | 0.161 | 0.252 |
| | Left Hippocampus | 0.038 | 0.013 | 2.990 | **0.003** | **0.010** |
| | Right Hippocampus | 0.030 | 0.013 | 2.406 | **0.016** | **0.040** |
| | WMH | -0.012 | 0.012 | -0.991 | 0.322 | 0.435 |

Significant results are highlighted in bold. False discovery rate (FDR) correction was applied across all brain measures and MHT variables listed in this table. Abbreviations: MRI = magnetic resonance imaging, S.E. = standard error, GM = grey matter, BAG = brain age gap, WM = white matter, WMH = white matter hyperintensity.

**Table S5| Associations between menopausal hormone therapy (MHT)-related variables and brain measures in the prescription MHT sample.**

| MHT Variable | MRI Measure | beta | S.E. | t-value | p-value | pFDR-value |
|---|---|---|---|---|---|---|
| **MHT formulation** | | | | | | |
| Estrogens-only | GM BAG | -0.025 | 0.068 | -0.367 | 0.714 | 0.994 |
| | WM BAG | 0.040 | 0.068 | 0.591 | 0.554 | 0.994 |
| | Left Hippocampus | -0.066 | 0.064 | -1.032 | 0.302 | 0.994 |
| | Right Hippocampus | -0.050 | 0.064 | -0.775 | 0.438 | 0.994 |
| | WMH | 0.017 | 0.060 | 0.283 | 0.777 | 0.994 |
| Estrogens+Progestin | GM BAG | 0.005 | 0.057 | 0.093 | 0.926 | 0.994 |
| | WM BAG | 0.127 | 0.057 | 2.221 | **0.026** | 0.832 |
| | Left Hippocampus | -0.010 | 0.054 | -0.179 | 0.858 | 0.994 |
| | Right Hippocampus | -0.011 | 0.054 | -0.198 | 0.843 | 0.994 |
| | WMH | 0.052 | 0.050 | 1.039 | 0.299 | 0.994 |
| **Route of Administration** | | | | | | |
| oral | GM BAG | -0.048 | 0.066 | -0.729 | 0.466 | 0.994 |
| | WM BAG | 0.113 | 0.066 | 1.708 | 0.088 | 0.994 |
| | Left Hippocampus | -0.030 | 0.063 | -0.478 | 0.632 | 0.994 |
| | Right Hippocampus | -0.001 | 0.063 | -0.023 | 0.981 | 0.994 |
| | WMH | 0.134 | 0.057 | 2.336 | **0.019** | 0.832 |
| transdermal | GM BAG | -0.205 | 0.125 | -1.636 | 0.102 | 0.994 |
| | WM BAG | -0.038 | 0.126 | -0.300 | 0.764 | 0.994 |
| | Left Hippocampus | -0.040 | 0.118 | -0.338 | 0.735 | 0.994 |
| | Right Hippocampus | -0.053 | 0.118 | -0.448 | 0.654 | 0.994 |
| | WMH | -0.094 | 0.110 | -0.852 | 0.394 | 0.994 |
| vaginal | GM BAG | 0.138 | 0.096 | 1.432 | 0.152 | 0.994 |
| | WM BAG | 0.143 | 0.096 | 1.485 | 0.138 | 0.994 |
| | Left Hippocampus | -0.052 | 0.091 | -0.571 | 0.568 | 0.994 |
| | Right Hippocampus | -0.002 | 0.091 | -0.022 | 0.983 | 0.994 |
| | WMH | 0.070 | 0.086 | 0.818 | 0.414 | 0.994 |
| injection | GM BAG | 0.446 | 0.333 | 1.338 | 0.181 | 0.994 |
| | WM BAG | 0.204 | 0.333 | 0.612 | 0.541 | 0.994 |
| | Left Hippocampus | -0.276 | 0.315 | -0.876 | 0.381 | 0.994 |
| | Right Hippocampus | 0.102 | 0.315 | 0.323 | 0.747 | 0.994 |
| | WMH | -0.072 | 0.286 | -0.251 | 0.802 | 0.994 |
| mixed | GM BAG | 0.012 | 0.091 | 0.128 | 0.898 | 0.994 |
| | WM BAG | 0.062 | 0.091 | 0.680 | 0.496 | 0.994 |
| | Left Hippocampus | -0.001 | 0.086 | -0.007 | 0.994 | 0.994 |
| | Right Hippocampus | -0.093 | 0.086 | -1.082 | 0.279 | 0.994 |
| | WMH | -0.101 | 0.079 | -1.274 | 0.203 | 0.994 |
| **Estrogen-only Forms** | | | | | | |
| Bioidentical | GM BAG | -0.013 | 0.076 | -0.169 | 0.866 | 0.994 |
| | WM BAG | 0.043 | 0.076 | 0.560 | 0.575 | 0.994 |
| | Left Hippocampus | -0.041 | 0.072 | -0.563 | 0.573 | 0.994 |
| | Right Hippocampus | -0.013 | 0.072 | -0.180 | 0.857 | 0.994 |
| | WMH | 0.001 | 0.067 | 0.012 | 0.991 | 0.994 |
| Synthetic | GM BAG | 0.053 | 0.214 | 0.248 | 0.804 | 0.994 |
| | WM BAG | 0.028 | 0.214 | 0.133 | 0.894 | 0.994 |
| | Left Hippocampus | -0.125 | 0.202 | -0.622 | 0.534 | 0.994 |
| | Right Hippocampus | -0.066 | 0.202 | -0.326 | 0.744 | 0.994 |
| | WMH | 0.106 | 0.184 | 0.580 | 0.562 | 0.994 |
| **Estrogen-only, active ingredient** | | | | | | |
| estradiol | GM BAG | -0.231 | 0.172 | -1.343 | 0.179 | 0.994 |
| | WM BAG | -0.133 | 0.172 | -0.771 | 0.441 | 0.994 |
| | Left Hippocampus | -0.049 | 0.162 | -0.303 | 0.762 | 0.994 |
| | Right Hippocampus | 0.020 | 0.162 | 0.121 | 0.904 | 0.994 |
| | WMH | -0.050 | 0.150 | -0.336 | 0.737 | 0.994 |
| estradiol hemihydrate | GM BAG | 0.039 | 0.085 | 0.465 | 0.642 | 0.994 |
| | WM BAG | 0.085 | 0.085 | 1.004 | 0.315 | 0.994 |

| | | | | | | |
|---|---|---|---|---|---|---|
| | Left Hippocampus | -0.038 | 0.080 | -0.479 | 0.632 | 0.994 |
| | Right Hippocampus | -0.021 | 0.080 | -0.258 | 0.796 | 0.994 |
| | WMH | 0.013 | 0.075 | 0.178 | 0.859 | 0.994 |
| estradiol valerate | GM BAG | 0.478 | 0.500 | 0.957 | 0.338 | 0.994 |
| | WM BAG | 0.259 | 0.500 | 0.517 | 0.605 | 0.994 |
| | Left Hippocampus | -0.024 | 0.472 | -0.051 | 0.960 | 0.994 |
| | Right Hippocampus | 0.388 | 0.472 | 0.823 | 0.410 | 0.994 |
| | WMH | 0.188 | 0.429 | 0.438 | 0.662 | 0.994 |
| CEE | GM BAG | -0.042 | 0.236 | -0.179 | 0.858 | 0.994 |
| | WM BAG | -0.022 | 0.236 | -0.095 | 0.925 | 0.994 |
| | Left Hippocampus | -0.148 | 0.223 | -0.662 | 0.508 | 0.994 |
| | Right Hippocampus | -0.167 | 0.223 | -0.747 | 0.455 | 0.994 |
| | WMH | 0.088 | 0.203 | 0.433 | 0.665 | 0.994 |
| Mixed | GM BAG | -0.177 | 0.193 | -0.914 | 0.361 | 0.994 |
| | WM BAG | 0.030 | 0.193 | 0.153 | 0.878 | 0.994 |
| | Left Hippocampus | -0.181 | 0.182 | -0.995 | 0.320 | 0.994 |
| | Right Hippocampus | -0.269 | 0.182 | -1.479 | 0.139 | 0.994 |
| | WMH | 0.039 | 0.169 | 0.232 | 0.816 | 0.994 |
| **Estrogens-only, Dosage (mg)** | | | | | | |
| | GM BAG | -0.045 | 0.074 | -0.605 | 0.546 | 0.994 |
| | WM BAG | -0.080 | 0.075 | -1.065 | 0.288 | 0.994 |
| | Left Hippocampus | -0.004 | 0.071 | -0.051 | 0.960 | 0.994 |
| | Right Hippocampus | -0.018 | 0.071 | -0.256 | 0.798 | 0.994 |
| | WMH | -0.007 | 0.061 | -0.108 | 0.914 | 0.994 |
| **Estrogens-only, Duration of Use (weeks)** | | | | | | |
| | GM BAG | -0.047 | 0.081 | -0.578 | 0.564 | 0.994 |
| | WM BAG | -0.141 | 0.085 | -1.648 | 0.101 | 0.994 |
| | Left Hippocampus | -0.007 | 0.083 | -0.090 | 0.928 | 0.994 |
| | Right Hippocampus | -0.037 | 0.079 | -0.475 | 0.635 | 0.994 |
| | WMH | -0.146 | 0.066 | -2.221 | **0.028** | 0.832 |
| **Estrogens + Progestins Form** | | | | | | |
| Bioidentical | GM BAG | -0.119 | 0.249 | -0.477 | 0.633 | 0.994 |
| | WM BAG | 0.011 | 0.250 | 0.044 | 0.965 | 0.994 |
| | Left Hippocampus | 0.108 | 0.236 | 0.457 | 0.647 | 0.994 |
| | Right Hippocampus | 0.336 | 0.236 | 1.424 | 0.154 | 0.994 |
| | WMH | 0.034 | 0.215 | 0.157 | 0.875 | 0.994 |
| Synthetic | GM BAG | -0.025 | 0.186 | -0.133 | 0.895 | 0.994 |
| | WM BAG | 0.204 | 0.186 | 1.093 | 0.275 | 0.994 |
| | Left Hippocampus | 0.134 | 0.176 | 0.760 | 0.447 | 0.994 |
| | Right Hippocampus | 0.162 | 0.176 | 0.921 | 0.357 | 0.994 |
| | WMH | 0.135 | 0.160 | 0.844 | 0.399 | 0.994 |
| Bioidentical & Synthetic | GM BAG | 0.009 | 0.105 | 0.089 | 0.929 | 0.994 |
| | WM BAG | 0.057 | 0.105 | 0.544 | 0.587 | 0.994 |
| | Left Hippocampus | -0.097 | 0.099 | -0.979 | 0.327 | 0.994 |
| | Right Hippocampus | -0.115 | 0.099 | -1.158 | 0.247 | 0.994 |
| | WMH | 0.075 | 0.091 | 0.819 | 0.413 | 0.994 |
| **Estrogens + Progestins, active ingredient** | | | | | | |
| estradiol hemihydrate & norethisterone acetate | GM BAG | -0.067 | 0.140 | -0.481 | 0.631 | 0.994 |
| | WM BAG | -0.022 | 0.140 | -0.155 | 0.877 | 0.994 |
| | Left Hippocampus | 0.045 | 0.132 | 0.338 | 0.735 | 0.994 |
| | Right Hippocampus | -0.005 | 0.132 | -0.041 | 0.967 | 0.994 |
| | WMH | 0.062 | 0.122 | 0.509 | 0.610 | 0.994 |
| estradiol hemihydrate & dydrogesterone | GM BAG | -0.200 | 0.267 | -0.749 | 0.454 | 0.994 |
| | WM BAG | -0.093 | 0.267 | -0.348 | 0.728 | 0.994 |
| | Left Hippocampus | 0.036 | 0.252 | 0.144 | 0.886 | 0.994 |

| | | | | | | |
|---|---|---|---|---|---|---|
| | Right Hippocampus | 0.321 | 0.252 | 1.271 | 0.204 | 0.994 |
| | WMH | -0.009 | 0.229 | -0.040 | 0.968 | 0.994 |
| estradiol hemihydrate & norethisterone | GM BAG | 0.277 | 0.277 | 1.001 | 0.317 | 0.994 |
| | WM BAG | 0.115 | 0.277 | 0.414 | 0.679 | 0.994 |
| | Left Hippocampus | -0.368 | 0.262 | -1.406 | 0.160 | 0.994 |
| | Right Hippocampus | -0.443 | 0.262 | -1.693 | 0.091 | 0.994 |
| | WMH | 0.354 | 0.238 | 1.488 | 0.137 | 0.994 |
| CEE & norgestrel | GM BAG | -0.076 | 0.229 | -0.331 | 0.741 | 0.994 |
| | WM BAG | -0.026 | 0.230 | -0.113 | 0.910 | 0.994 |
| | Left Hippocampus | 0.134 | 0.217 | 0.619 | 0.536 | 0.994 |
| | Right Hippocampus | 0.036 | 0.217 | 0.164 | 0.869 | 0.994 |
| | WMH | 0.136 | 0.197 | 0.689 | 0.491 | 0.994 |
| CEE & medroxyprogesterone acetate | GM BAG | 0.074 | 0.316 | 0.234 | 0.815 | 0.994 |
| | WM BAG | 0.634 | 0.316 | 2.004 | **0.045** | 0.994 |
| | Left Hippocampus | 0.128 | 0.298 | 0.429 | 0.668 | 0.994 |
| | Right Hippocampus | 0.399 | 0.298 | 1.336 | 0.182 | 0.994 |
| | WMH | 0.130 | 0.271 | 0.481 | 0.631 | 0.994 |
| tibolone | GM BAG | -0.441 | 0.277 | -1.592 | 0.111 | 0.994 |
| | WM BAG | 0.025 | 0.278 | 0.090 | 0.929 | 0.994 |
| | Left Hippocampus | 0.033 | 0.262 | 0.126 | 0.900 | 0.994 |
| | Right Hippocampus | 0.166 | 0.262 | 0.633 | 0.526 | 0.994 |
| | WMH | -0.180 | 0.248 | -0.725 | 0.469 | 0.994 |
| Mixed | GM BAG | 0.067 | 0.079 | 0.854 | 0.393 | 0.994 |
| | WM BAG | 0.179 | 0.079 | 2.270 | **0.023** | 0.832 |
| | Left Hippocampus | -0.002 | 0.075 | -0.030 | 0.976 | 0.994 |
| | Right Hippocampus | -0.035 | 0.075 | -0.476 | 0.634 | 0.994 |
| | WMH | 0.037 | 0.069 | 0.536 | 0.592 | 0.994 |
| **Estrogens + Progestins, Progestin Generation** | | | | | | |
| 1stGen | GM BAG | 0.054 | 0.094 | 0.579 | 0.563 | 0.994 |
| | WM BAG | 0.139 | 0.094 | 1.485 | 0.138 | 0.994 |
| | Left Hippocampus | -0.044 | 0.088 | -0.499 | 0.618 | 0.994 |
| | Right Hippocampus | -0.039 | 0.088 | -0.439 | 0.661 | 0.994 |
| | WMH | 0.088 | 0.082 | 1.079 | 0.281 | 0.994 |
| 2ndGen | GM BAG | -0.004 | 0.156 | -0.025 | 0.980 | 0.994 |
| | WM BAG | 0.068 | 0.157 | 0.432 | 0.666 | 0.994 |
| | Left Hippocampus | 0.083 | 0.148 | 0.564 | 0.573 | 0.994 |
| | Right Hippocampus | 0.105 | 0.148 | 0.714 | 0.475 | 0.994 |
| | WMH | 0.119 | 0.135 | 0.883 | 0.377 | 0.994 |
| **Estrogens + Progestins, Dosage (mg)** | | | | | | |
| Estrogens | GM BAG | -0.068 | 0.058 | -1.185 | 0.237 | 0.994 |
| | WM BAG | -0.004 | 0.058 | -0.077 | 0.938 | 0.994 |
| | Left Hippocampus | -0.053 | 0.056 | -0.950 | 0.343 | 0.994 |
| | Right Hippocampus | 0.001 | 0.056 | 0.016 | 0.987 | 0.994 |
| | WMH | 0.063 | 0.053 | 1.192 | 0.234 | 0.994 |
| Progestins | GM BAG | 0.062 | 0.057 | 1.099 | 0.273 | 0.994 |
| | WM BAG | 0.039 | 0.057 | 0.684 | 0.495 | 0.994 |
| | Left Hippocampus | 0.072 | 0.055 | 1.292 | 0.197 | 0.994 |
| | Right Hippocampus | 0.055 | 0.055 | 0.984 | 0.326 | 0.994 |
| | WMH | 0.071 | 0.051 | 1.390 | 0.166 | 0.994 |
| **Estrogens + Progestins, Duration of Use (weeks)** | | | | | | |
| Estrogens | GM BAG | 0.089 | 0.071 | 1.247 | 0.213 | 0.994 |
| | WM BAG | -0.022 | 0.073 | -0.297 | 0.767 | 0.994 |
| | Left Hippocampus | -0.149 | 0.069 | -2.165 | **0.031** | 0.832 |
| | Right Hippocampus | -0.165 | 0.068 | -2.438 | **0.015** | 0.832 |
| | WMH | -0.093 | 0.066 | -1.417 | 0.158 | 0.994 |

| | | | | | | |
|---|---|---|---|---|---|---|
| Progestins | GM BAG | -0.015 | 0.074 | -0.207 | 0.836 | 0.994 |
| | WM BAG | 0.071 | 0.076 | 0.938 | 0.349 | 0.994 |
| | Left Hippocampus | -0.026 | 0.072 | -0.368 | 0.713 | 0.994 |
| | Right Hippocampus | 0.021 | 0.071 | 0.295 | 0.768 | 0.994 |
| | WMH | 0.045 | 0.068 | 0.654 | 0.514 | 0.994 |

Significant results are highlighted in bold. False discovery rate (FDR) correction was applied across all brain measures and MHT variables listed in this table. Abbreviations: MRI = magnetic resonance imaging, S.E. = standard error, GM = grey matter, BAG = brain age gap, WM = white matter, WMH = white matter hyperintensity, CEE = conjugated equine estrogen, Gen = generation.

**Table S6| Associations between APOE ε4 genotype and brain measures in the whole sample.**

| Genotype | MRI Measure | beta | S.E. | t-value | p-value | pFDR-value |
|---|---|---|---|---|---|---|
| APOE ε4 | GM BAG | 0.017 | 0.008 | 2.060 | **0.039** | 0.118 |
| ε3/ε4 | GM BAG | 0.040 | 0.019 | 2.070 | **0.038** | 0.118 |
| ε4/ε4 | GM BAG | 0.022 | 0.053 | 0.422 | 0.673 | 0.721 |
| APOE ε4 | WM BAG | 0.011 | 0.008 | 1.372 | 0.170 | 0.252 |
| ε3/ε4 | WM BAG | 0.027 | 0.019 | 1.382 | 0.167 | 0.252 |
| ε4/ε4 | WM BAG | 0.014 | 0.053 | 0.270 | 0.787 | 0.787 |
| APOE ε4 | Left Hippocampus | -0.019 | 0.008 | -2.467 | **0.014** | 0.068 |
| ε3/ε4 | Left Hippocampus | -0.034 | 0.018 | -1.865 | 0.062 | 0.138 |
| ε4/ε4 | Left Hippocampus | -0.134 | 0.050 | -2.699 | **0.007** | 0.052 |
| APOE ε4 | Right Hippocampus | -0.010 | 0.008 | -1.327 | 0.185 | 0.252 |
| ε3/ε4 | Right Hippocampus | -0.010 | 0.018 | -0.531 | 0.595 | 0.687 |
| ε4/ε4 | Right Hippocampus | -0.156 | 0.050 | -3.135 | **0.002** | **0.026** |
| APOE ε4 | WMH | 0.013 | 0.007 | 1.849 | 0.064 | 0.138 |
| ε3/ε4 | WMH | 0.027 | 0.017 | 1.640 | 0.101 | 0.189 |
| ε4/ε4 | WMH | 0.053 | 0.046 | 1.154 | 0.249 | 0.311 |

Significant results are highlighted in bold. False discovery rate (FDR) correction was applied across all brain measures and MHT variables listed in this table. Abbreviations: APOE = apolipoprotein, MRI = magnetic resonance imaging, S.E. = standard error, GM = grey matter, BAG = brain age gap, WM = white matter, WMH = white matter hyperintensity.

**Table S7| Interactions between APOE ε4 genotype and menopausal hormone therapy (MHT)-related variables on brain measures in the entire sample.**

| MHT Variable | MRI Measure | beta | S.E. | t-value | p-value | pFDR-value |
|---|---|---|---|---|---|---|
| **MHT Status * APOE ε4** | GM BAG | 0.009 | 0.008 | 1.050 | 0.294 | 0.927 |
| | WM BAG | 0.007 | 0.008 | 0.796 | 0.426 | 0.927 |
| | Left Hippocampus | -0.008 | 0.008 | -1.089 | 0.276 | 0.927 |
| | Right Hippocampus | -0.010 | 0.008 | -1.344 | 0.179 | 0.927 |
| | WMH | -0.001 | 0.007 | -0.150 | 0.880 | 0.945 |
| **Current MHT use * APOE ε4** | GM BAG | 0.047 | 0.040 | 1.189 | 0.234 | 0.927 |
| | WM BAG | 0.014 | 0.040 | 0.363 | 0.717 | 0.927 |
| | Left Hippocampus | 0.029 | 0.037 | 0.772 | 0.440 | 0.927 |
| | Right Hippocampus | 0.047 | 0.037 | 1.275 | 0.202 | 0.927 |
| | WMH | -0.006 | 0.034 | -0.170 | 0.865 | 0.945 |
| **Past MHT use * APOE ε4** | GM BAG | 0.013 | 0.020 | 0.637 | 0.524 | 0.927 |
| | WM BAG | 0.006 | 0.020 | 0.281 | 0.778 | 0.927 |
| | Left Hippocampus | -0.031 | 0.019 | -1.639 | 0.101 | 0.927 |
| | Right Hippocampus | -0.037 | 0.019 | -1.963 | 0.050 | 0.927 |
| | WMH | -0.001 | 0.018 | -0.083 | 0.934 | 0.945 |
| **Age at first MHT use * APOE ε4** | GM BAG | 0.018 | 0.016 | 1.088 | 0.277 | 0.927 |
| | WM BAG | 0.006 | 0.016 | 0.377 | 0.706 | 0.927 |
| | Left Hippocampus | 0.013 | 0.015 | 0.813 | 0.416 | 0.927 |
| | Right Hippocampus | 0.008 | 0.015 | 0.534 | 0.593 | 0.927 |
| | WMH | 0.006 | 0.014 | 0.441 | 0.660 | 0.927 |
| **Age at first MHT use relative to age at menopause * APOE ε4** | GM BAG | -0.017 | 0.019 | -0.899 | 0.369 | 0.927 |
| | WM BAG | 0.006 | 0.019 | 0.331 | 0.741 | 0.927 |
| | Left Hippocampus | -0.004 | 0.018 | -0.207 | 0.836 | 0.945 |
| | Right Hippocampus | -0.006 | 0.018 | -0.344 | 0.731 | 0.927 |
| | WMH | -0.026 | 0.016 | -1.604 | 0.109 | 0.927 |
| **Age at last MHT use * APOE** | GM BAG | 0.015 | 0.018 | 0.814 | 0.416 | 0.927 |
| | WM BAG | -0.007 | 0.018 | -0.397 | 0.691 | 0.927 |
| | Left Hippocampus | 0.010 | 0.018 | 0.558 | 0.577 | 0.927 |
| | Right Hippocampus | -0.002 | 0.017 | -0.092 | 0.927 | 0.945 |
| | WMH | -0.022 | 0.016 | -1.383 | 0.167 | 0.927 |
| **Age at last MHT use relative to age at menopause * APOE ε4** | GM BAG | -0.007 | 0.017 | -0.437 | 0.662 | 0.927 |
| | WM BAG | 0.015 | 0.017 | 0.868 | 0.386 | 0.927 |
| | Left Hippocampus | -0.003 | 0.016 | -0.202 | 0.840 | 0.945 |
| | Right Hippocampus | -0.006 | 0.016 | -0.386 | 0.699 | 0.927 |
| | WMH | -0.008 | 0.014 | -0.561 | 0.575 | 0.927 |
| **Duration of MHT use * APOE ε4** | GM BAG | 0.006 | 0.015 | 0.359 | 0.720 | 0.927 |
| | WM BAG | -0.008 | 0.015 | -0.534 | 0.594 | 0.927 |
| | Left Hippocampus | -0.010 | 0.014 | -0.690 | 0.490 | 0.927 |
| | Right Hippocampus | -0.013 | 0.014 | -0.910 | 0.363 | 0.927 |
| | WMH | -0.016 | 0.013 | -1.172 | 0.241 | 0.927 |
| **Bilateral Oophorectomy * APOE ε4** | GM BAG | -0.001 | 0.012 | -0.069 | 0.945 | 0.945 |
| | WM BAG | 0.019 | 0.012 | 1.639 | 0.101 | 0.927 |
| | Left Hippocampus | 0.006 | 0.011 | 0.581 | 0.561 | 0.927 |
| | Right Hippocampus | 0.005 | 0.011 | 0.493 | 0.622 | 0.927 |
| | WMH | 0.013 | 0.010 | 1.239 | 0.216 | 0.927 |
| **Hysterectomy * APOE ε4** | GM BAG | -0.004 | 0.014 | -0.291 | 0.771 | 0.927 |
| | WM BAG | -0.001 | 0.014 | -0.074 | 0.941 | 0.945 |
| | Left Hippocampus | -0.004 | 0.013 | -0.310 | 0.756 | 0.927 |
| | Right Hippocampus | -0.014 | 0.013 | -1.043 | 0.297 | 0.927 |
| | WMH | -0.025 | 0.012 | -1.995 | **0.046** | 0.927 |

Significant results are highlighted in bold. False discovery rate (FDR) correction was applied across all brain measures and MHT variables listed in this table. Abbreviations: APOE = apolipoprotein, MRI = magnetic resonance imaging, S.E. = standard error, GM = grey matter, BAG = brain age gap, WM = white matter, WMH = white matter hyperintensity.

**Table S8| Interactions between APOE ε4 genotype and menopausal hormone therapy (MHT)-related variables on brain measures in the prescription sample.**

| MHT Variable | MRI Measure | beta | S.E. | t-value | p-value | pFDR-value |
|---|---|---|---|---|---|---|
| **MHT formulation * APOE ε4** | | | | | | |
| Estrogens-only | GM BAG | 0.072 | 0.067 | 1.076 | 0.282 | 0.649 |
| | WM BAG | 0.077 | 0.067 | 1.150 | 0.250 | 0.649 |
| | Left Hippocampus | -0.010 | 0.063 | -0.152 | 0.879 | 0.976 |
| | Right Hippocampus | -0.095 | 0.063 | -1.501 | 0.133 | 0.649 |
| | WMH | -0.016 | 0.060 | -0.273 | 0.785 | 0.941 |
| Estrogens+Progestin | GM BAG | -0.032 | 0.060 | -0.535 | 0.592 | 0.872 |
| | WM BAG | -0.033 | 0.060 | -0.545 | 0.586 | 0.872 |
| | Left Hippocampus | 0.008 | 0.057 | 0.143 | 0.887 | 0.976 |
| | Right Hippocampus | -0.057 | 0.057 | -0.998 | 0.319 | 0.681 |
| | WMH | -0.072 | 0.052 | -1.397 | 0.163 | 0.649 |
| **Route of Administration * APOE ε4** | | | | | | |
| oral | GM BAG | -0.008 | 0.068 | -0.115 | 0.908 | 0.976 |
| | WM BAG | 0.059 | 0.068 | 0.872 | 0.383 | 0.730 |
| | Left Hippocampus | 0.003 | 0.064 | 0.039 | 0.969 | 0.981 |
| | Right Hippocampus | -0.063 | 0.064 | -0.992 | 0.321 | 0.681 |
| | WMH | -0.056 | 0.058 | -0.958 | 0.338 | 0.685 |
| transdermal | GM BAG | 0.077 | 0.132 | 0.579 | 0.563 | 0.865 |
| | WM BAG | -0.104 | 0.132 | -0.784 | 0.433 | 0.778 |
| | Left Hippocampus | 0.170 | 0.125 | 1.362 | 0.173 | 0.649 |
| | Right Hippocampus | 0.039 | 0.125 | 0.309 | 0.757 | 0.941 |
| | WMH | -0.139 | 0.114 | -1.219 | 0.223 | 0.649 |
| vaginal | GM BAG | 0.096 | 0.091 | 1.048 | 0.295 | 0.649 |
| | WM BAG | 0.015 | 0.091 | 0.159 | 0.873 | 0.976 |
| | Left Hippocampus | -0.004 | 0.086 | -0.043 | 0.965 | 0.981 |
| | Right Hippocampus | -0.109 | 0.086 | -1.267 | 0.205 | 0.649 |
| | WMH | 0.002 | 0.083 | 0.023 | 0.981 | 0.981 |
| injection | GM BAG | 0.558 | 0.326 | 1.712 | 0.087 | 0.649 |
| | WM BAG | 0.106 | 0.326 | 0.327 | 0.744 | 0.941 |
| | Left Hippocampus | -0.703 | 0.307 | -2.289 | **0.022** | 0.449 |
| | Right Hippocampus | -0.462 | 0.307 | -1.503 | 0.133 | 0.649 |
| | WMH | -0.114 | 0.280 | -0.408 | 0.683 | 0.934 |
| mixed | GM BAG | -0.181 | 0.104 | -1.736 | 0.083 | 0.649 |
| | WM BAG | -0.067 | 0.104 | -0.639 | 0.523 | 0.837 |
| | Left Hippocampus | -0.027 | 0.098 | -0.278 | 0.781 | 0.941 |
| | Right Hippocampus | -0.114 | 0.098 | -1.162 | 0.245 | 0.649 |
| | WMH | -0.067 | 0.090 | -0.743 | 0.458 | 0.796 |
| **Estrogen-only Forms * APOE ε4** | | | | | | |
| Bioidentical | GM BAG | 0.092 | 0.075 | 1.229 | 0.219 | 0.649 |
| | WM BAG | 0.032 | 0.075 | 0.422 | 0.673 | 0.928 |
| | Left Hippocampus | 0.038 | 0.071 | 0.536 | 0.592 | 0.872 |
| | Right Hippocampus | -0.080 | 0.071 | -1.133 | 0.257 | 0.649 |
| | WMH | -0.040 | 0.067 | -0.601 | 0.548 | 0.857 |
| Synthetic | GM BAG | -0.223 | 0.197 | -1.132 | 0.258 | 0.649 |
| | WM BAG | 0.271 | 0.197 | 1.373 | 0.170 | 0.649 |
| | Left Hippocampus | -0.149 | 0.186 | -0.803 | 0.422 | 0.767 |
| | Right Hippocampus | -0.012 | 0.186 | -0.064 | 0.949 | 0.981 |
| | WMH | -0.155 | 0.169 | -0.916 | 0.360 | 0.693 |
| **Estrogen-only, active ingredient * APOE ε4** | | | | | | |
| estradiol | GM BAG | 0.079 | 0.181 | 0.435 | 0.664 | 0.927 |

| | | | | | | |
|---|---|---|---|---|---|---|
| | WM BAG | -0.126 | 0.181 | -0.698 | 0.485 | 0.824 |
| | Left Hippocampus | 0.113 | 0.170 | 0.663 | 0.508 | 0.831 |
| | Right Hippocampus | 0.043 | 0.171 | 0.250 | 0.803 | 0.941 |
| | WMH | -0.148 | 0.156 | -0.947 | 0.344 | 0.687 |
| estradiol hemihydrate | GM BAG | 0.087 | 0.083 | 1.052 | 0.293 | 0.649 |
| | WM BAG | 0.057 | 0.083 | 0.692 | 0.489 | 0.824 |
| | Left Hippocampus | 0.022 | 0.078 | 0.280 | 0.779 | 0.941 |
| | Right Hippocampus | -0.105 | 0.078 | -1.352 | 0.177 | 0.649 |
| | WMH | -0.018 | 0.075 | -0.240 | 0.811 | 0.941 |
| estradiol valerate | GM BAG | -0.228 | 0.516 | -0.443 | 0.658 | 0.927 |
| | WM BAG | 0.985 | 0.516 | 1.911 | 0.056 | 0.561 |
| | Left Hippocampus | -0.818 | 0.486 | -1.684 | 0.092 | 0.649 |
| | Right Hippocampus | -0.537 | 0.486 | -1.104 | 0.270 | 0.649 |
| | WMH | -0.155 | 0.443 | -0.350 | 0.726 | 0.941 |
| CEE | GM BAG | -0.332 | 0.227 | -1.465 | 0.143 | 0.649 |
| | WM BAG | 0.120 | 0.227 | 0.530 | 0.596 | 0.872 |
| | Left Hippocampus | -0.057 | 0.214 | -0.267 | 0.789 | 0.941 |
| | Right Hippocampus | 0.010 | 0.214 | 0.047 | 0.963 | 0.981 |
| | WMH | -0.191 | 0.195 | -0.979 | 0.328 | 0.681 |
| Mixed | GM BAG | 0.234 | 0.243 | 0.962 | 0.336 | 0.685 |
| | WM BAG | 0.272 | 0.243 | 1.121 | 0.262 | 0.649 |
| | Left Hippocampus | -0.327 | 0.229 | -1.428 | 0.153 | 0.649 |
| | Right Hippocampus | -0.491 | 0.229 | -2.145 | **0.032** | 0.511 |
| | WMH | 0.400 | 0.209 | 1.911 | 0.056 | 0.561 |
| **Estrogens-only, Dosage (mg) * APOE ε4** | | | | | | |
| | GM BAG | -0.019 | 0.072 | -0.266 | 0.791 | 0.941 |
| | WM BAG | 0.078 | 0.072 | 1.091 | 0.276 | 0.649 |
| | Left Hippocampus | -0.073 | 0.068 | -1.064 | 0.288 | 0.649 |
| | Right Hippocampus | -0.003 | 0.068 | -0.045 | 0.964 | 0.981 |
| | WMH | -0.010 | 0.059 | -0.175 | 0.862 | 0.976 |
| **Estrogens-only, Duration of Use (weeks) * APOE ε4** | | | | | | |
| | GM BAG | -0.046 | 0.077 | -0.596 | 0.552 | 0.857 |
| | WM BAG | 0.019 | 0.080 | 0.242 | 0.809 | 0.941 |
| | Left Hippocampus | -0.052 | 0.078 | -0.661 | 0.510 | 0.831 |
| | Right Hippocampus | -0.002 | 0.074 | -0.024 | 0.981 | 0.981 |
| | WMH | -0.010 | 0.062 | -0.165 | 0.869 | 0.976 |
| **Estrogens + Progestins Form * APOE ε4** | | | | | | |
| Bioidentical | GM BAG | -0.067 | 0.257 | -0.263 | 0.793 | 0.941 |
| | WM BAG | -0.327 | 0.258 | -1.268 | 0.205 | 0.649 |
| | Left Hippocampus | -0.349 | 0.243 | -1.437 | 0.151 | 0.649 |
| | Right Hippocampus | -0.341 | 0.243 | -1.402 | 0.161 | 0.649 |
| | WMH | 0.303 | 0.221 | 1.370 | 0.171 | 0.649 |
| Synthetic | GM BAG | -0.512 | 0.220 | -2.330 | **0.020** | 0.449 |
| | WM BAG | -0.323 | 0.220 | -1.464 | 0.143 | 0.649 |
| | Left Hippocampus | 0.502 | 0.207 | 2.420 | **0.016** | 0.449 |
| | Right Hippocampus | 0.405 | 0.208 | 1.947 | 0.052 | 0.561 |
| | WMH | -0.412 | 0.189 | -2.180 | **0.029** | 0.511 |
| Bioidentical & Synthetic | GM BAG | 0.106 | 0.114 | 0.931 | 0.352 | 0.693 |
| | WM BAG | 0.096 | 0.114 | 0.840 | 0.401 | 0.746 |
| | Left Hippocampus | -0.135 | 0.108 | -1.251 | 0.211 | 0.649 |
| | Right Hippocampus | -0.292 | 0.108 | -2.709 | **0.007** | 0.449 |
| | WMH | -0.061 | 0.098 | -0.621 | 0.535 | 0.847 |
| **Estrogens + Progestins, active ingredient * APOE ε4** | | | | | | |
| estradiol hemihydrate & norethisterone acetate | GM BAG | 0.247 | 0.148 | 1.662 | 0.097 | 0.649 |

| | | | | | | |
|---|---|---|---|---|---|---|
| | WM BAG | 0.137 | 0.149 | 0.925 | 0.355 | 0.693 |
| | Left Hippocampus | -0.219 | 0.140 | -1.565 | 0.118 | 0.649 |
| | Right Hippocampus | -0.320 | 0.140 | -2.283 | **0.022** | 0.449 |
| | WMH | -0.209 | 0.128 | -1.635 | 0.102 | 0.649 |
| estradiol hemihydrate & dydrogesterone | GM BAG | -0.020 | 0.264 | -0.076 | 0.940 | 0.981 |
| | WM BAG | -0.279 | 0.264 | -1.056 | 0.291 | 0.649 |
| | Left Hippocampus | -0.322 | 0.249 | -1.296 | 0.195 | 0.649 |
| | Right Hippocampus | -0.349 | 0.249 | -1.400 | 0.162 | 0.649 |
| | WMH | 0.345 | 0.226 | 1.525 | 0.127 | 0.649 |
| estradiol hemihydrate & norethisterone | GM BAG | -0.382 | 0.293 | -1.303 | 0.193 | 0.649 |
| | WM BAG | -0.154 | 0.294 | -0.525 | 0.599 | 0.872 |
| | Left Hippocampus | 0.030 | 0.277 | 0.109 | 0.914 | 0.976 |
| | Right Hippocampus | -0.324 | 0.277 | -1.169 | 0.242 | 0.649 |
| | WMH | 0.124 | 0.252 | 0.492 | 0.622 | 0.889 |
| CEE & norgestrel | GM BAG | -0.513 | 0.253 | -2.029 | **0.042** | 0.561 |
| | WM BAG | -0.348 | 0.253 | -1.373 | 0.170 | 0.649 |
| | Left Hippocampus | 0.414 | 0.238 | 1.737 | 0.082 | 0.649 |
| | Right Hippocampus | 0.437 | 0.239 | 1.830 | 0.067 | 0.633 |
| | WMH | -0.499 | 0.217 | -2.299 | **0.022** | 0.449 |
| CEE & medroxyprogesterone acetate | GM BAG | -0.512 | 0.470 | -1.090 | 0.276 | 0.649 |
| | WM BAG | -0.050 | 0.470 | -0.106 | 0.915 | 0.976 |
| | Left Hippocampus | 0.860 | 0.443 | 1.941 | 0.052 | 0.561 |
| | Right Hippocampus | 0.493 | 0.444 | 1.111 | 0.266 | 0.649 |
| | WMH | -0.159 | 0.403 | -0.394 | 0.693 | 0.939 |
| tibolone | GM BAG | -0.709 | 0.464 | -1.527 | 0.127 | 0.649 |
| | WM BAG | -0.401 | 0.465 | -0.862 | 0.389 | 0.731 |
| | Left Hippocampus | 0.881 | 0.437 | 2.013 | **0.044** | 0.561 |
| | Right Hippocampus | 0.468 | 0.438 | 1.068 | 0.285 | 0.649 |
| | WMH | -0.418 | 0.400 | -1.045 | 0.296 | 0.649 |
| Mixed | GM BAG | -0.039 | 0.079 | -0.493 | 0.622 | 0.889 |
| | WM BAG | -0.026 | 0.079 | -0.328 | 0.743 | 0.941 |
| | Left Hippocampus | 0.024 | 0.075 | 0.324 | 0.746 | 0.941 |
| | Right Hippocampus | 0.018 | 0.075 | 0.238 | 0.812 | 0.941 |
| | WMH | -0.057 | 0.068 | -0.831 | 0.406 | 0.746 |
| **Estrogens + Progestins, Progestin Generation * APOE ε4** | | | | | | |
| 1stGen | GM BAG | 0.113 | 0.103 | 1.096 | 0.273 | 0.649 |
| | WM BAG | 0.040 | 0.103 | 0.387 | 0.699 | 0.939 |
| | Left Hippocampus | -0.134 | 0.097 | -1.374 | 0.169 | 0.649 |
| | Right Hippocampus | -0.231 | 0.097 | -2.370 | **0.018** | 0.449 |
| | WMH | -0.104 | 0.089 | -1.162 | 0.245 | 0.649 |
| 2ndGen | GM BAG | -0.239 | 0.169 | -1.413 | 0.158 | 0.649 |
| | WM BAG | -0.250 | 0.169 | -1.476 | 0.140 | 0.649 |
| | Left Hippocampus | 0.024 | 0.159 | 0.148 | 0.882 | 0.976 |
| | Right Hippocampus | 0.044 | 0.160 | 0.275 | 0.783 | 0.941 |
| | WMH | -0.113 | 0.145 | -0.775 | 0.438 | 0.779 |
| **Estrogens + Progestins, Dosage (mg) * APOE ε4** | | | | | | |
| Estrogens | GM BAG | 0.070 | 0.061 | 1.148 | 0.252 | 0.649 |
| | WM BAG | 0.076 | 0.061 | 1.248 | 0.213 | 0.649 |
| | Left Hippocampus | -0.008 | 0.059 | -0.128 | 0.899 | 0.976 |
| | Right Hippocampus | -0.058 | 0.059 | -0.988 | 0.324 | 0.681 |
| | WMH | -0.024 | 0.054 | -0.432 | 0.666 | 0.927 |
| Progestins | GM BAG | -0.061 | 0.221 | -0.276 | 0.783 | 0.941 |
| | WM BAG | -0.160 | 0.222 | -0.724 | 0.470 | 0.808 |
| | Left Hippocampus | -0.059 | 0.216 | -0.275 | 0.783 | 0.941 |

| | | | | | | |
|---|---|---|---|---|---|---|
| | Right Hippocampus | 0.113 | 0.214 | 0.528 | 0.598 | 0.872 |
| | WMH | 0.512 | 0.197 | 2.591 | **0.010** | 0.449 |
| **Estrogens + Progestins, Duration of Use (weeks) * APOE ε4** | | | | | | |
| Estrogens | GM BAG | 0.013 | 0.070 | 0.190 | 0.850 | 0.976 |
| | WM BAG | -0.047 | 0.072 | -0.653 | 0.514 | 0.831 |
| | Left Hippocampus | -0.005 | 0.068 | -0.067 | 0.946 | 0.981 |
| | Right Hippocampus | 0.043 | 0.066 | 0.655 | 0.513 | 0.831 |
| | WMH | 0.070 | 0.064 | 1.106 | 0.270 | 0.649 |
| Progestins | GM BAG | -0.025 | 0.080 | -0.317 | 0.751 | 0.941 |
| | WM BAG | 0.101 | 0.081 | 1.243 | 0.215 | 0.649 |
| | Left Hippocampus | 0.059 | 0.077 | 0.763 | 0.446 | 0.784 |
| | Right Hippocampus | -0.007 | 0.075 | -0.095 | 0.924 | 0.979 |
| | WMH | 0.009 | 0.072 | 0.122 | 0.903 | 0.976 |

Significant results are highlighted in bold. False discovery rate (FDR) correction was applied across all brain measures and MHT variables listed in this table. Abbreviations: APOE = apolipoprotein, MRI = magnetic resonance imaging, S.E. = standard error, GM = grey matter, BAG = brain age gap, WM = white matter, WMH = white matter hyperintensity, CEE = conjugated equine estrogen, Gen = generation.

**Table S9| Associations between menopausal hormone therapy (MHT)-related variables and brain measures in the whole sample, excluding participants with ICD-10 diagnosis known to impact the brain.**

| MHT Variable | MRI Measure | beta | S.E. | t-value | p-value | pFDR-value |
|---|---|---|---|---|---|---|
| MHT Status | GM BAG | 0.036 | 0.009 | 4.155 | **3.27e-05** | **0.001** |
| | WM BAG | 0.017 | 0.009 | 1.952 | 0.051 | 0.109 |
| | Left Hippocampus | -0.021 | 0.008 | -2.615 | **0.009** | **0.032** |
| | Right Hippocampus | -0.012 | 0.008 | -1.462 | 0.144 | 0.257 |
| | WMH | 0.007 | 0.007 | 0.890 | 0.374 | 0.505 |
| Current MHT use | GM BAG | 0.224 | 0.040 | 5.644 | **1.70e-08** | **8.49e-07** |
| | WM BAG | 0.160 | 0.040 | 4.042 | **5.33e-05** | **0.001** |
| | Left Hippocampus | -0.158 | 0.037 | -4.257 | **2.08e-05** | **0.001** |
| | Right Hippocampus | -0.130 | 0.037 | -3.531 | **4.16e-04** | **0.003** |
| | WMH | -0.007 | 0.034 | -0.203 | 0.839 | 0.933 |
| Past MHT use | GM BAG | 0.046 | 0.022 | 2.111 | **0.035** | 0.092 |
| | WM BAG | 0.003 | 0.022 | 0.135 | 0.892 | 0.949 |
| | Left Hippocampus | -0.011 | 0.020 | -0.552 | 0.581 | 0.764 |
| | Right Hippocampus | -0.002 | 0.020 | -0.108 | 0.914 | 0.952 |
| | WMH | 0.020 | 0.018 | 1.077 | 0.281 | 0.426 |
| Age at first MHT use | GM BAG | 0.004 | 0.016 | 0.257 | 0.798 | 0.906 |
| | WM BAG | 0.000 | 0.016 | -0.007 | 0.995 | 0.995 |
| | Left Hippocampus | 0.004 | 0.016 | 0.262 | 0.793 | 0.906 |
| | Right Hippocampus | -0.006 | 0.016 | -0.405 | 0.685 | 0.857 |
| | WMH | -0.022 | 0.014 | -1.585 | 0.113 | 0.209 |
| Age at first MHT use relative to age at menopause | GM BAG | 0.024 | 0.017 | 1.388 | 0.165 | 0.275 |
| | WM BAG | 0.036 | 0.017 | 2.083 | **0.037** | 0.092 |
| | Left Hippocampus | 0.009 | 0.017 | 0.524 | 0.600 | 0.770 |
| | Right Hippocampus | -0.017 | 0.017 | -1.018 | 0.309 | 0.441 |
| | WMH | -0.004 | 0.015 | -0.298 | 0.765 | 0.906 |
| Age at last MHT use | GM BAG | 0.039 | 0.019 | 2.069 | **0.039** | 0.092 |
| | WM BAG | 0.032 | 0.019 | 1.662 | 0.097 | 0.192 |
| | Left Hippocampus | -0.030 | 0.018 | -1.646 | 0.100 | 0.192 |
| | Right Hippocampus | -0.022 | 0.018 | -1.187 | 0.235 | 0.368 |
| | WMH | 0.023 | 0.017 | 1.364 | 0.173 | 0.278 |
| Age at last MHT use relative to age at menopause | GM BAG | 0.053 | 0.019 | 2.798 | **0.005** | **0.022** |
| | WM BAG | 0.062 | 0.019 | 3.254 | **0.001** | **0.006** |
| | Left Hippocampus | -0.039 | 0.019 | -2.089 | **0.037** | 0.092 |
| | Right Hippocampus | -0.038 | 0.019 | -2.052 | **0.040** | 0.092 |
| | WMH | 0.043 | 0.017 | 2.588 | **0.010** | **0.032** |
| Duration of MHT use | GM BAG | 0.064 | 0.017 | 3.787 | 1.55e-04 | **0.002** |
| | WM BAG | 0.054 | 0.017 | 3.192 | **0.001** | **0.007** |
| | Left Hippocampus | -0.055 | 0.016 | -3.465 | **0.001** | **0.003** |
| | Right Hippocampus | -0.035 | 0.016 | -2.235 | **0.025** | 0.075 |
| | WMH | 0.028 | 0.015 | 1.942 | 0.052 | 0.109 |
| Bilateral Oophorectomy | GM BAG | -0.033 | 0.013 | -2.594 | **0.009** | **0.032** |
| | WM BAG | -0.004 | 0.013 | -0.291 | 0.771 | 0.906 |
| | Left Hippocampus | 0.012 | 0.012 | 1.046 | 0.296 | 0.435 |
| | Right Hippocampus | -0.001 | 0.012 | -0.076 | 0.939 | 0.958 |
| | WMH | 0.002 | 0.011 | 0.164 | 0.869 | 0.945 |
| Hysterectomy | GM BAG | -0.047 | 0.013 | -3.497 | **4.74e-04** | **0.003** |
| | WM BAG | -0.019 | 0.014 | -1.401 | 0.161 | 0.275 |
| | Left Hippocampus | 0.038 | 0.013 | 2.990 | **0.003** | **0.013** |
| | Right Hippocampus | 0.030 | 0.013 | 2.406 | **0.016** | 0.051 |
| | WMH | -0.012 | 0.012 | -0.991 | 0.322 | 0.447 |

Significant results are highlighted in bold. False discovery rate (FDR) correction was applied across all brain measures and MHT variables listed in this table. Abbreviations: MRI = magnetic resonance imaging, S.E. = standard error, GM = grey matter, BAG = brain age gap, WM = white matter, WMH = white matter hyperintensity.

**Table S11| Associations between menopausal hormone therapy (MHT)-related variables and brain measures in the prescription MHT sample, excluding participants with ICD-10 diagnosis known to impact the brain.**

| MHT Variable | MRI Measure | beta | S.E. | t-value | p-value | pFDR-value |
|---|---|---|---|---|---|---|
| **MHT formulation** | | | | | | |
| Estrogens-only | GM BAG | -0.008 | 0.071 | -0.118 | 0.906 | 0.990 |
| | WM BAG | 0.044 | 0.071 | 0.625 | 0.532 | 0.990 |
| | Left Hippocampus | -0.054 | 0.067 | -0.809 | 0.419 | 0.990 |
| | Right Hippocampus | -0.058 | 0.067 | -0.861 | 0.389 | 0.990 |
| | WMH | 0.021 | 0.062 | 0.333 | 0.739 | 0.990 |
| Estrogens+Progestin | GM BAG | -0.045 | 0.060 | -0.739 | 0.460 | 0.990 |
| | WM BAG | 0.080 | 0.060 | 1.332 | 0.183 | 0.990 |
| | Left Hippocampus | 0.025 | 0.057 | 0.439 | 0.661 | 0.990 |
| | Right Hippocampus | 0.023 | 0.057 | 0.407 | 0.684 | 0.990 |
| | WMH | 0.007 | 0.052 | 0.138 | 0.890 | 0.990 |
| **Route of Administration** | | | | | | |
| oral | GM BAG | -0.081 | 0.070 | -1.163 | 0.245 | 0.990 |
| | WM BAG | 0.079 | 0.070 | 1.138 | 0.255 | 0.990 |
| | Left Hippocampus | 0.011 | 0.066 | 0.160 | 0.873 | 0.990 |
| | Right Hippocampus | 0.017 | 0.066 | 0.257 | 0.797 | 0.990 |
| | WMH | 0.103 | 0.060 | 1.711 | 0.087 | 0.990 |
| transdermal | GM BAG | -0.248 | 0.134 | -1.848 | 0.065 | 0.990 |
| | WM BAG | -0.086 | 0.134 | -0.643 | 0.520 | 0.990 |
| | Left Hippocampus | -0.023 | 0.126 | -0.183 | 0.855 | 0.990 |
| | Right Hippocampus | -0.046 | 0.126 | -0.368 | 0.713 | 0.990 |
| | WMH | -0.117 | 0.115 | -1.011 | 0.312 | 0.990 |
| vaginal | GM BAG | 0.168 | 0.099 | 1.696 | 0.090 | 0.990 |
| | WM BAG | 0.146 | 0.099 | 1.470 | 0.142 | 0.990 |
| | Left Hippocampus | -0.036 | 0.093 | -0.389 | 0.697 | 0.990 |
| | Right Hippocampus | -0.023 | 0.093 | -0.251 | 0.802 | 0.990 |
| | WMH | 0.092 | 0.088 | 1.050 | 0.294 | 0.990 |
| injection | GM BAG | 0.536 | 0.353 | 1.515 | 0.130 | 0.990 |
| | WM BAG | 0.290 | 0.354 | 0.820 | 0.412 | 0.990 |
| | Left Hippocampus | -0.173 | 0.333 | -0.518 | 0.605 | 0.990 |
| | Right Hippocampus | 0.182 | 0.333 | 0.547 | 0.584 | 0.990 |
| | WMH | -0.054 | 0.302 | -0.181 | 0.857 | 0.990 |
| mixed | GM BAG | -0.046 | 0.096 | -0.481 | 0.630 | 0.990 |
| | WM BAG | 0.024 | 0.096 | 0.249 | 0.804 | 0.990 |
| | Left Hippocampus | 0.003 | 0.090 | 0.031 | 0.976 | 0.990 |
| | Right Hippocampus | -0.047 | 0.090 | -0.516 | 0.606 | 0.990 |
| | WMH | -0.161 | 0.083 | -1.940 | 0.052 | 0.990 |
| **Estrogen-only Forms** | | | | | | |
| Bioidentical | GM BAG | 0.012 | 0.080 | 0.151 | 0.880 | 0.990 |
| | WM BAG | 0.048 | 0.080 | 0.602 | 0.547 | 0.990 |
| | Left Hippocampus | -0.039 | 0.075 | -0.518 | 0.604 | 0.990 |
| | Right Hippocampus | -0.031 | 0.075 | -0.417 | 0.677 | 0.990 |
| | WMH | 0.008 | 0.070 | 0.117 | 0.907 | 0.990 |
| Synthetic | GM BAG | 0.146 | 0.230 | 0.636 | 0.525 | 0.990 |
| | WM BAG | 0.105 | 0.230 | 0.455 | 0.649 | 0.990 |
| | Left Hippocampus | -0.100 | 0.217 | -0.462 | 0.644 | 0.990 |
| | Right Hippocampus | -0.005 | 0.216 | -0.025 | 0.980 | 0.990 |
| | WMH | 0.063 | 0.196 | 0.319 | 0.750 | 0.990 |
| **Estrogen-only, active ingredient** | | | | | | |
| estradiol | GM BAG | -0.243 | 0.186 | -1.306 | 0.192 | 0.990 |
| | WM BAG | -0.174 | 0.186 | -0.935 | 0.350 | 0.990 |
| | Left Hippocampus | -0.108 | 0.175 | -0.614 | 0.539 | 0.990 |
| | Right Hippocampus | -0.009 | 0.175 | -0.049 | 0.961 | 0.990 |
| | WMH | -0.123 | 0.162 | -0.759 | 0.448 | 0.990 |

| | | | | | | |
|---|---|---|---|---|---|---|
| estradiol hemihydrate | GM BAG | 0.069 | 0.088 | 0.779 | 0.436 | 0.990 |
| | WM BAG | 0.098 | 0.088 | 1.109 | 0.268 | 0.990 |
| | Left Hippocampus | -0.024 | 0.083 | -0.284 | 0.776 | 0.990 |
| | Right Hippocampus | -0.036 | 0.083 | -0.438 | 0.661 | 0.990 |
| | WMH | 0.038 | 0.078 | 0.487 | 0.626 | 0.990 |
| estradiol valerate | GM BAG | 0.482 | 0.500 | 0.965 | 0.335 | 0.990 |
| | WM BAG | 0.258 | 0.500 | 0.515 | 0.606 | 0.990 |
| | Left Hippocampus | -0.030 | 0.471 | -0.064 | 0.949 | 0.990 |
| | Right Hippocampus | 0.386 | 0.470 | 0.822 | 0.411 | 0.990 |
| | WMH | 0.203 | 0.426 | 0.476 | 0.634 | 0.990 |
| CEE | GM BAG | 0.056 | 0.259 | 0.215 | 0.829 | 0.990 |
| | WM BAG | 0.064 | 0.259 | 0.247 | 0.805 | 0.990 |
| | Left Hippocampus | -0.118 | 0.244 | -0.486 | 0.627 | 0.990 |
| | Right Hippocampus | -0.110 | 0.243 | -0.450 | 0.653 | 0.990 |
| | WMH | 0.024 | 0.221 | 0.110 | 0.912 | 0.990 |
| Mixed | GM BAG | -0.255 | 0.197 | -1.294 | 0.196 | 0.990 |
| | WM BAG | -0.027 | 0.197 | -0.136 | 0.892 | 0.990 |
| | Left Hippocampus | -0.114 | 0.185 | -0.616 | 0.538 | 0.990 |
| | Right Hippocampus | -0.255 | 0.185 | -1.374 | 0.169 | 0.990 |
| | WMH | 0.058 | 0.168 | 0.348 | 0.728 | 0.990 |
| **Estrogens-only, Dosage (mg)** | | | | | | |
| | GM BAG | -0.047 | 0.078 | -0.603 | 0.547 | 0.990 |
| | WM BAG | -0.097 | 0.079 | -1.232 | 0.219 | 0.990 |
| | Left Hippocampus | -0.031 | 0.075 | -0.411 | 0.681 | 0.990 |
| | Right Hippocampus | -0.041 | 0.074 | -0.550 | 0.583 | 0.990 |
| | WMH | -0.026 | 0.064 | -0.399 | 0.690 | 0.990 |
| **Estrogens-only, Duration of Use (weeks)** | | | | | | |
| | GM BAG | -0.101 | 0.086 | -1.171 | 0.244 | 0.990 |
| | WM BAG | -0.131 | 0.092 | -1.431 | 0.155 | 0.990 |
| | Left Hippocampus | -0.003 | 0.089 | -0.033 | 0.974 | 0.990 |
| | Right Hippocampus | 0.003 | 0.084 | 0.041 | 0.968 | 0.990 |
| | WMH | -0.158 | 0.070 | -2.248 | **0.026** | 0.990 |
| **Estrogens + Progestins Form** | | | | | | |
| Bioidentical | GM BAG | -0.248 | 0.258 | -0.962 | 0.336 | 0.990 |
| | WM BAG | 0.000 | 0.259 | 0.000 | 1.000 | 1.000 |
| | Left Hippocampus | 0.091 | 0.243 | 0.372 | 0.710 | 0.990 |
| | Right Hippocampus | 0.388 | 0.243 | 1.595 | 0.111 | 0.990 |
| | WMH | -0.011 | 0.221 | -0.049 | 0.961 | 0.990 |
| Synthetic | GM BAG | -0.099 | 0.196 | -0.506 | 0.613 | 0.990 |
| | WM BAG | 0.133 | 0.197 | 0.678 | 0.498 | 0.990 |
| | Left Hippocampus | 0.182 | 0.185 | 0.980 | 0.327 | 0.990 |
| | Right Hippocampus | 0.153 | 0.185 | 0.826 | 0.409 | 0.990 |
| | WMH | 0.170 | 0.168 | 1.012 | 0.312 | 0.990 |
| Bioidentical & Synthetic | GM BAG | -0.063 | 0.111 | -0.564 | 0.573 | 0.990 |
| | WM BAG | -0.026 | 0.112 | -0.236 | 0.813 | 0.990 |
| | Left Hippocampus | -0.049 | 0.105 | -0.463 | 0.644 | 0.990 |
| | Right Hippocampus | -0.071 | 0.105 | -0.674 | 0.500 | 0.990 |
| | WMH | 0.002 | 0.096 | 0.021 | 0.983 | 0.990 |
| **Estrogens + Progestins, active ingredient** | | | | | | |
| estradiol hemihydrate & norethisterone acetate | GM BAG | -0.176 | 0.151 | -1.167 | 0.243 | 0.990 |
| | WM BAG | -0.103 | 0.151 | -0.680 | 0.497 | 0.990 |
| | Left Hippocampus | 0.147 | 0.142 | 1.034 | 0.301 | 0.990 |
| | Right Hippocampus | 0.078 | 0.142 | 0.547 | 0.585 | 0.990 |
| | WMH | -0.011 | 0.130 | -0.086 | 0.932 | 0.990 |
| estradiol hemihydrate & dydrogesterone | GM BAG | -0.355 | 0.277 | -1.283 | 0.200 | 0.990 |

| | | | | | | |
|---|---|---|---|---|---|---|
| | WM BAG | -0.114 | 0.277 | -0.412 | 0.680 | 0.990 |
| | Left Hippocampus | 0.012 | 0.261 | 0.047 | 0.962 | 0.990 |
| | Right Hippocampus | 0.380 | 0.261 | 1.455 | 0.146 | 0.990 |
| | WMH | -0.068 | 0.237 | -0.286 | 0.775 | 0.990 |
| estradiol hemihydrate & norethisterone | GM BAG | 0.276 | 0.301 | 0.916 | 0.360 | 0.990 |
| | WM BAG | -0.153 | 0.302 | -0.509 | 0.611 | 0.990 |
| | Left Hippocampus | -0.332 | 0.284 | -1.168 | 0.243 | 0.990 |
| | Right Hippocampus | -0.345 | 0.284 | -1.216 | 0.224 | 0.990 |
| | WMH | 0.147 | 0.257 | 0.570 | 0.569 | 0.990 |
| CEE & norgestrel | GM BAG | -0.077 | 0.229 | -0.336 | 0.737 | 0.990 |
| | WM BAG | -0.029 | 0.230 | -0.127 | 0.899 | 0.990 |
| | Left Hippocampus | 0.129 | 0.217 | 0.597 | 0.551 | 0.990 |
| | Right Hippocampus | 0.033 | 0.216 | 0.154 | 0.878 | 0.990 |
| | WMH | 0.146 | 0.196 | 0.743 | 0.458 | 0.990 |
| CEE & medroxyprogesterone acetate | GM BAG | -0.159 | 0.377 | -0.421 | 0.674 | 0.990 |
| | WM BAG | 0.566 | 0.378 | 1.497 | 0.134 | 0.990 |
| | Left Hippocampus | 0.319 | 0.356 | 0.896 | 0.370 | 0.990 |
| | Right Hippocampus | 0.475 | 0.355 | 1.337 | 0.181 | 0.990 |
| | WMH | 0.231 | 0.322 | 0.718 | 0.473 | 0.990 |
| tibolone | GM BAG | -0.444 | 0.277 | -1.603 | 0.109 | 0.990 |
| | WM BAG | 0.020 | 0.278 | 0.072 | 0.943 | 0.990 |
| | Left Hippocampus | 0.028 | 0.262 | 0.109 | 0.913 | 0.990 |
| | Right Hippocampus | 0.166 | 0.261 | 0.635 | 0.525 | 0.990 |
| | WMH | -0.173 | 0.246 | -0.701 | 0.483 | 0.990 |
| Mixed | GM BAG | 0.043 | 0.083 | 0.522 | 0.602 | 0.990 |
| | WM BAG | 0.151 | 0.083 | 1.819 | 0.069 | 0.990 |
| | Left Hippocampus | 0.025 | 0.078 | 0.321 | 0.748 | 0.990 |
| | Right Hippocampus | -0.005 | 0.078 | -0.068 | 0.946 | 0.990 |
| | WMH | 0.009 | 0.072 | 0.129 | 0.898 | 0.990 |
| **Estrogens + Progestins, Progestin Generation** | | | | | | |
| 1stGen | GM BAG | -0.032 | 0.101 | -0.315 | 0.753 | 0.990 |
| | WM BAG | 0.053 | 0.101 | 0.529 | 0.597 | 0.990 |
| | Left Hippocampus | 0.003 | 0.095 | 0.030 | 0.976 | 0.990 |
| | Right Hippocampus | 0.008 | 0.095 | 0.086 | 0.932 | 0.990 |
| | WMH | 0.003 | 0.087 | 0.035 | 0.972 | 0.990 |
| 2ndGen | GM BAG | -0.071 | 0.160 | -0.440 | 0.660 | 0.990 |
| | WM BAG | 0.022 | 0.161 | 0.136 | 0.892 | 0.990 |
| | Left Hippocampus | 0.055 | 0.151 | 0.362 | 0.717 | 0.990 |
| | Right Hippocampus | 0.101 | 0.151 | 0.669 | 0.504 | 0.990 |
| | WMH | 0.086 | 0.137 | 0.624 | 0.532 | 0.990 |
| **Estrogens + Progestins, Dosage (mg)** | | | | | | |
| Estrogens | GM BAG | -0.098 | 0.060 | -1.625 | 0.105 | 0.990 |
| | WM BAG | 0.009 | 0.061 | 0.149 | 0.882 | 0.990 |
| | Left Hippocampus | -0.017 | 0.059 | -0.286 | 0.775 | 0.990 |
| | Right Hippocampus | 0.045 | 0.059 | 0.767 | 0.443 | 0.990 |
| | WMH | 0.058 | 0.055 | 1.054 | 0.293 | 0.990 |
| Progestins | GM BAG | 0.041 | 0.060 | 0.686 | 0.493 | 0.990 |
| | WM BAG | 0.040 | 0.060 | 0.671 | 0.503 | 0.990 |
| | Left Hippocampus | 0.079 | 0.058 | 1.368 | 0.173 | 0.990 |
| | Right Hippocampus | 0.082 | 0.058 | 1.427 | 0.155 | 0.990 |
| | WMH | 0.057 | 0.053 | 1.088 | 0.278 | 0.990 |
| **Estrogens + Progestins, Duration of Use (weeks)** | | | | | | |
| Estrogens | GM BAG | 0.091 | 0.074 | 1.232 | 0.219 | 0.990 |
| | WM BAG | -0.010 | 0.076 | -0.129 | 0.898 | 0.990 |
| | Left Hippocampus | -0.172 | 0.071 | -2.405 | **0.017** | 0.990 |

| | | | | | | |
|---|---|---|---|---|---|---|
| | Right Hippocampus | -0.177 | 0.070 | -2.522 | **0.012** | 0.990 |
| | WMH | -0.089 | 0.067 | -1.317 | 0.189 | 0.990 |
| Progestins | GM BAG | -0.003 | 0.078 | -0.034 | 0.973 | 0.990 |
| | WM BAG | 0.062 | 0.080 | 0.775 | 0.439 | 0.990 |
| | Left Hippocampus | -0.017 | 0.075 | -0.221 | 0.826 | 0.990 |
| | Right Hippocampus | 0.030 | 0.074 | 0.407 | 0.684 | 0.990 |
| | WMH | 0.023 | 0.071 | 0.330 | 0.742 | 0.990 |

Significant results are highlighted in bold. False discovery rate (FDR) correction was applied across all brain measures and MHT variables listed in this table. Abbreviations: MRI = magnetic resonance imaging, S.E. = standard error, GM = grey matter, BAG = brain age gap, WM = white matter, WMH = white matter hyperintensity, CEE = conjugated equine estrogen, Gen = generation.

**Table S11| Associations between menopausal hormone therapy (MHT)-related variables and brain measures in the whole sample, also adjusting for age$^2$.**

| MHT Variable | MRI Measure | beta | S.E. | t-value | p-value | pFDR-value |
|---|---|---|---|---|---|---|
| MHT Status | BAG GM | 0.029 | 0.008 | 3.513 | **4.44e-04** | **0.003** |
| | BAG WM | 0.015 | 0.008 | 1.844 | 0.065 | 0.136 |
| | Left Hippocampus | -0.017 | 0.008 | -2.236 | **0.025** | 0.064 |
| | Right Hippocampus | -0.007 | 0.008 | -0.921 | 0.357 | 0.497 |
| | WMH | 0.007 | 0.007 | 0.918 | 0.359 | 0.497 |
| Current MHT use | BAG GM | 0.217 | 0.037 | 5.798 | **6.84e-09** | **3.42e-07** |
| | BAG WM | 0.154 | 0.038 | 4.099 | **4.18e-05** | **4.18e-04** |
| | Left Hippocampus | -0.152 | 0.035 | -4.349 | **1.38e-05** | **1.72e-04** |
| | Right Hippocampus | -0.129 | 0.035 | -3.678 | **2.36e-04** | **0.002** |
| | WMH | -0.005 | 0.032 | -0.161 | 0.872 | 0.948 |
| Past MHT use | BAG GM | 0.026 | 0.021 | 1.268 | 0.205 | 0.353 |
| | BAG WM | 0.001 | 0.021 | 0.069 | 0.945 | 0.982 |
| | Left Hippocampus | -0.001 | 0.019 | -0.048 | 0.962 | 0.982 |
| | Right Hippocampus | 0.014 | 0.019 | 0.715 | 0.475 | 0.597 |
| | WMH | 0.019 | 0.018 | 1.048 | 0.295 | 0.460 |
| Age at first MHT use | BAG GM | 0.003 | 0.016 | 0.213 | 0.832 | 0.945 |
| | BAG WM | 0.000 | 0.015 | -0.018 | 0.986 | 0.986 |
| | Left Hippocampus | 0.010 | 0.015 | 0.685 | 0.493 | 0.602 |
| | Right Hippocampus | -0.001 | 0.015 | -0.055 | 0.956 | 0.982 |
| | WMH | -0.024 | 0.013 | -1.792 | 0.073 | 0.147 |
| Age at first MHT use relative to age at menopause | BAG GM | 0.018 | 0.016 | 1.092 | 0.275 | 0.443 |
| | BAG WM | 0.023 | 0.016 | 1.388 | 0.165 | 0.306 |
| | Left Hippocampus | 0.013 | 0.016 | 0.795 | 0.427 | 0.562 |
| | Right Hippocampus | -0.016 | 0.016 | -1.027 | 0.304 | 0.461 |
| | WMH | -0.010 | 0.014 | -0.711 | 0.477 | 0.597 |
| Age at last MHT use | BAG GM | 0.044 | 0.018 | 2.448 | **0.014** | **0.042** |
| | BAG WM | 0.037 | 0.018 | 2.075 | **0.038** | 0.091 |
| | Left Hippocampus | -0.033 | 0.017 | -1.901 | 0.057 | 0.125 |
| | Right Hippocampus | -0.024 | 0.017 | -1.408 | 0.159 | 0.306 |
| | WMH | 0.018 | 0.016 | 1.155 | 0.248 | 0.414 |
| Age at last MHT use relative to age at menopause | BAG GM | 0.056 | 0.018 | 3.060 | **0.002** | **0.009** |
| | BAG WM | 0.057 | 0.018 | 3.150 | **0.002** | **0.008** |
| | Left Hippocampus | -0.045 | 0.018 | -2.564 | **0.010** | **0.035** |
| | Right Hippocampus | -0.045 | 0.017 | -2.600 | **0.009** | **0.033** |
| | WMH | 0.035 | 0.016 | 2.231 | **0.026** | 0.064 |
| Duration of MHT use | BAG GM | 0.074 | 0.016 | 4.601 | **4.33e-06** | **1.08e-04** |
| | BAG WM | 0.064 | 0.016 | 3.964 | **7.51e-05** | **0.001** |
| | Left Hippocampus | -0.067 | 0.015 | -4.474 | **7.89e-06** | **1.32e-04** |
| | Right Hippocampus | -0.046 | 0.015 | -3.107 | **0.002** | **0.009** |
| | WMH | 0.028 | 0.014 | 1.968 | **0.049** | 0.112 |
| Bilateral Oophorectomy | BAG GM | -0.031 | 0.013 | -2.476 | **0.013** | **0.042** |
| | BAG WM | -0.002 | 0.013 | -0.189 | 0.850 | 0.945 |
| | Left Hippocampus | 0.011 | 0.012 | 0.901 | 0.368 | 0.497 |
| | Right Hippocampus | -0.003 | 0.012 | -0.256 | 0.798 | 0.945 |
| | WMH | 0.002 | 0.011 | 0.221 | 0.825 | 0.945 |
| Hysterectomy | BAG GM | -0.046 | 0.013 | -3.409 | **0.001** | **0.004** |
| | BAG WM | -0.018 | 0.013 | -1.325 | 0.185 | 0.331 |
| | Left Hippocampus | 0.036 | 0.013 | 2.885 | **0.004** | **0.015** |
| | Right Hippocampus | 0.028 | 0.012 | 2.279 | **0.023** | 0.063 |
| | WMH | -0.011 | 0.012 | -0.951 | 0.342 | 0.497 |

Significant results are highlighted in bold. False discovery rate (FDR) correction was applied across all brain measures and MHT variables listed in this table. Abbreviations: MRI = magnetic resonance imaging, S.E. = standard error, GM = grey matter, BAG = brain age gap, WM = white matter, WMH = white matter hyperintensity.

**Table S12| Associations between menopausal hormone therapy (MHT)-related variables and brain measures in the whole sample, after removal of extreme values.**

| MHT Variable | MRI Measure | beta | S.E. | t-value | p-value | pFDR-value |
|---|---|---|---|---|---|---|
| Age at first MHT use | GM BAG | 0.001 | 0.016 | 0.059 | 0.953 | 0.953 |
| | WM BAG | -0.005 | 0.016 | -0.300 | 0.764 | 0.882 |
| | Left Hippocampus | 0.004 | 0.015 | 0.285 | 0.776 | 0.882 |
| | Right Hippocampus | -0.010 | 0.015 | -0.659 | 0.510 | 0.637 |
| | WMH | -0.028 | 0.014 | -2.041 | **0.041** | 0.086 |
| Age at first MHT use relative | GM BAG | 0.026 | 0.018 | 1.456 | 0.145 | 0.242 |
| to age at menopause | WM BAG | 0.041 | 0.018 | 2.333 | **0.020** | **0.049** |
| | Left Hippocampus | 0.002 | 0.017 | 0.132 | 0.895 | 0.933 |
| | Right Hippocampus | -0.038 | 0.017 | -2.207 | **0.027** | 0.062 |
| | WMH | -0.002 | 0.015 | -0.164 | 0.870 | 0.933 |
| Age at last MHT use | GM BAG | 0.050 | 0.018 | 2.743 | **0.006** | **0.026** |
| | WM BAG | 0.036 | 0.018 | 1.986 | 0.047 | 0.091 |
| | Left Hippocampus | -0.025 | 0.018 | -1.423 | 0.155 | 0.242 |
| | Right Hippocampus | -0.024 | 0.017 | -1.365 | 0.172 | 0.253 |
| | WMH | 0.011 | 0.016 | 0.727 | 0.467 | 0.614 |
| Age at last MHT use relative | GM BAG | 0.076 | 0.020 | 3.892 | **1.02e-04** | **0.001** |
| to age at menopause | WM BAG | 0.079 | 0.019 | 4.042 | **5.44e-05** | **0.001** |
| | Left Hippocampus | -0.048 | 0.019 | -2.511 | **0.012** | **0.038** |
| | Right Hippocampus | -0.056 | 0.019 | -2.938 | **0.003** | **0.024** |
| | WMH | 0.031 | 0.017 | 1.812 | 0.070 | 0.125 |
| Duration of MHT use | GM BAG | 0.051 | 0.018 | 2.819 | **0.005** | **0.024** |
| | WM BAG | 0.048 | 0.018 | 2.640 | **0.008** | **0.030** |
| | Left Hippocampus | -0.048 | 0.017 | -2.846 | **0.004** | **0.024** |
| | Right Hippocampus | -0.022 | 0.017 | -1.316 | 0.188 | 0.262 |
| | WMH | 0.039 | 0.016 | 2.431 | **0.015** | **0.042** |

Significant results are highlighted in bold. False discovery rate (FDR) correction was applied across all brain measures and MHT variables listed in this table. Abbreviations: MRI = magnetic resonance imaging, S.E. = standard error, GM = grey matter, BAG = brain age gap, WM = white matter, WMH = white matter hyperintensity.

**Table S13| Detected extreme values of continuous menopausal hormone therapy (MHT)-related variables using the median absolute deviation method.**

| | | | | Number of extreme values | | |
|---|---|---|---|---|---|---|
| **MHT Variable** | **Median** | **MAD** | **Limits*** | **Low** | **High** | **Total** |
| Age started MHT | 50 | 4.45 | 36.66 – 63.34 | 76 | 29 | 105 |
| Age last used MHT | 54 | 5.93 | 36.21 – 71.79 | 14 | 13 | 27 |
| Age at Menopause | 51 | 2.97 | 42.10 – 59.90 | 715 | 158 | 873 |

*Limits of acceptable range of values. Abbreviation: MHT = hormone therapy, MAD = median absolute deviation.

**Table S14| Associations between menopausal hormone therapy (MHT)-related variables and brain measures in the prescription MHT sample, adjusting for additional covariates.**

| MHT Variable | MRI Measure | beta | S.E. | t-value | p-value | pFDR-value |
|---|---|---|---|---|---|---|
| **MHT formulation** | | | | | | |
| Estrogens-only | GM BAG | -0.014 | 0.078 | -0.183 | 0.855 | 0.999 |
| | WM BAG | 0.048 | 0.078 | 0.613 | 0.540 | 0.999 |
| | Left Hippocampus | -0.070 | 0.073 | -0.955 | 0.339 | 0.999 |
| | Right Hippocampus | -0.060 | 0.073 | -0.811 | 0.418 | 0.999 |
| | WMH | 0.055 | 0.068 | 0.807 | 0.420 | 0.999 |
| Estrogens+Progestin | GM BAG | 0.011 | 0.060 | 0.182 | 0.856 | 0.999 |
| | WM BAG | 0.106 | 0.060 | 1.768 | 0.077 | 0.999 |
| | Left Hippocampus | 0.023 | 0.056 | 0.410 | 0.682 | 0.999 |
| | Right Hippocampus | 0.007 | 0.056 | 0.117 | 0.907 | 0.999 |
| | WMH | 0.045 | 0.052 | 0.875 | 0.382 | 0.999 |
| **Route of Administration** | | | | | | |
| oral | GM BAG | -0.060 | 0.071 | -0.856 | 0.392 | 0.999 |
| | WM BAG | 0.074 | 0.071 | 1.041 | 0.298 | 0.999 |
| | Left Hippocampus | -0.008 | 0.067 | -0.122 | 0.903 | 0.999 |
| | Right Hippocampus | 0.026 | 0.067 | 0.396 | 0.692 | 0.999 |
| | WMH | 0.121 | 0.061 | 1.984 | **0.047** | 0.986 |
| transdermal | GM BAG | -0.208 | 0.142 | -1.469 | 0.142 | 0.999 |
| | WM BAG | 0.033 | 0.142 | 0.229 | 0.819 | 0.999 |
| | Left Hippocampus | -0.029 | 0.134 | -0.215 | 0.830 | 0.999 |
| | Right Hippocampus | -0.100 | 0.134 | -0.747 | 0.455 | 0.999 |
| | WMH | -0.099 | 0.125 | -0.794 | 0.427 | 0.999 |
| vaginal | GM BAG | 0.097 | 0.103 | 0.941 | 0.347 | 0.999 |
| | WM BAG | 0.067 | 0.103 | 0.652 | 0.515 | 0.999 |
| | Left Hippocampus | -0.001 | 0.097 | -0.014 | 0.989 | 0.999 |
| | Right Hippocampus | 0.038 | 0.098 | 0.393 | 0.694 | 0.999 |
| | WMH | 0.072 | 0.091 | 0.782 | 0.434 | 0.999 |
| injection | GM BAG | 0.993 | 0.406 | 2.444 | **0.015** | 0.847 |
| | WM BAG | 0.374 | 0.408 | 0.917 | 0.359 | 0.999 |
| | Left Hippocampus | -0.660 | 0.384 | -1.717 | 0.086 | 0.999 |
| | Right Hippocampus | -0.396 | 0.385 | -1.029 | 0.303 | 0.999 |
| | WMH | -0.208 | 0.347 | -0.601 | 0.548 | 0.999 |
| mixed | GM BAG | 0.078 | 0.097 | 0.800 | 0.424 | 0.999 |
| | WM BAG | 0.128 | 0.098 | 1.309 | 0.191 | 0.999 |
| | Left Hippocampus | 0.019 | 0.092 | 0.202 | 0.840 | 0.999 |
| | Right Hippocampus | -0.094 | 0.092 | -1.021 | 0.307 | 0.999 |
| | WMH | -0.031 | 0.085 | -0.369 | 0.712 | 0.999 |
| **Estrogen-only Forms** | | | | | | |
| Bioidentical | GM BAG | 0.010 | 0.085 | 0.115 | 0.909 | 0.999 |
| | WM BAG | 0.030 | 0.086 | 0.354 | 0.723 | 0.999 |
| | Left Hippocampus | -0.034 | 0.081 | -0.420 | 0.674 | 0.999 |
| | Right Hippocampus | -0.020 | 0.081 | -0.249 | 0.804 | 0.999 |
| | WMH | 0.033 | 0.075 | 0.443 | 0.658 | 0.999 |
| Synthetic | GM BAG | 0.043 | 0.237 | 0.180 | 0.857 | 0.999 |
| | WM BAG | 0.025 | 0.237 | 0.106 | 0.916 | 0.999 |
| | Left Hippocampus | -0.216 | 0.224 | -0.964 | 0.335 | 0.999 |
| | Right Hippocampus | -0.075 | 0.224 | -0.332 | 0.740 | 0.999 |
| | WMH | 0.076 | 0.202 | 0.376 | 0.707 | 0.999 |
| **Estrogen-only, active ingredient** | | | | | | |
| estradiol | GM BAG | -0.127 | 0.196 | -0.646 | 0.518 | 0.999 |
| | WM BAG | 0.066 | 0.197 | 0.335 | 0.738 | 0.999 |
| | Left Hippocampus | -0.051 | 0.186 | -0.273 | 0.785 | 0.999 |
| | Right Hippocampus | -0.132 | 0.186 | -0.710 | 0.478 | 0.999 |
| | WMH | 0.121 | 0.171 | 0.706 | 0.480 | 0.999 |
| estradiol hemihydrate | GM BAG | 0.041 | 0.094 | 0.431 | 0.667 | 0.999 |
| | WM BAG | 0.022 | 0.095 | 0.238 | 0.812 | 0.999 |

| | | | | | | |
|---|---|---|---|---|---|---|
| | Left Hippocampus | -0.030 | 0.089 | -0.336 | 0.737 | 0.999 |
| | Right Hippocampus | 0.005 | 0.089 | 0.058 | 0.954 | 0.999 |
| | WMH | 0.013 | 0.084 | 0.158 | 0.875 | 0.999 |
| estradiol valerate | GM BAG | 0.442 | 0.498 | 0.888 | 0.375 | 0.999 |
| | WM BAG | 0.277 | 0.500 | 0.554 | 0.579 | 0.999 |
| | Left Hippocampus | -0.015 | 0.471 | -0.032 | 0.974 | 0.999 |
| | Right Hippocampus | 0.378 | 0.472 | 0.801 | 0.423 | 0.999 |
| | WMH | 0.174 | 0.425 | 0.409 | 0.682 | 0.999 |
| CEE | GM BAG | -0.074 | 0.268 | -0.277 | 0.782 | 0.999 |
| | WM BAG | -0.046 | 0.269 | -0.173 | 0.863 | 0.999 |
| | Left Hippocampus | -0.272 | 0.253 | -1.074 | 0.283 | 0.999 |
| | Right Hippocampus | -0.206 | 0.254 | -0.810 | 0.418 | 0.999 |
| | WMH | 0.050 | 0.229 | 0.217 | 0.828 | 0.999 |
| Mixed | GM BAG | -0.304 | 0.259 | -1.174 | 0.241 | 0.999 |
| | WM BAG | 0.260 | 0.260 | 1.003 | 0.316 | 0.999 |
| | Left Hippocampus | -0.223 | 0.245 | -0.910 | 0.363 | 0.999 |
| | Right Hippocampus | -0.394 | 0.245 | -1.605 | 0.109 | 0.999 |
| | WMH | 0.252 | 0.229 | 1.103 | 0.270 | 0.999 |
| **Estrogens-only, Dosage (mg)** | | | | | | |
| | GM BAG | 0.000 | 0.091 | 0.004 | 0.996 | 0.999 |
| | WM BAG | -0.042 | 0.090 | -0.462 | 0.645 | 0.999 |
| | Left Hippocampus | 0.001 | 0.084 | 0.014 | 0.989 | 0.999 |
| | Right Hippocampus | 0.001 | 0.084 | 0.016 | 0.987 | 0.999 |
| | WMH | 0.073 | 0.072 | 1.003 | 0.317 | 0.999 |
| **Estrogens-only, Duration of Use (weeks)** | | | | | | |
| | GM BAG | -0.067 | 0.100 | -0.670 | 0.504 | 0.999 |
| | WM BAG | -0.081 | 0.104 | -0.774 | 0.441 | 0.999 |
| | Left Hippocampus | -0.047 | 0.100 | -0.472 | 0.638 | 0.999 |
| | Right Hippocampus | -0.024 | 0.098 | -0.242 | 0.809 | 0.999 |
| | WMH | -0.154 | 0.078 | -1.989 | **0.049** | 0.986 |
| **Estrogens + Progestins Form** | | | | | | |
| Bioidentical | GM BAG | -0.253 | 0.256 | -0.985 | 0.325 | 0.999 |
| | WM BAG | -0.004 | 0.258 | -0.017 | 0.986 | 0.999 |
| | Left Hippocampus | 0.100 | 0.243 | 0.410 | 0.682 | 0.999 |
| | Right Hippocampus | 0.393 | 0.244 | 1.612 | 0.107 | 0.999 |
| | WMH | -0.044 | 0.220 | -0.199 | 0.842 | 0.999 |
| Synthetic | GM BAG | -0.166 | 0.199 | -0.834 | 0.404 | 0.999 |
| | WM BAG | -0.014 | 0.200 | -0.069 | 0.945 | 0.999 |
| | Left Hippocampus | 0.227 | 0.189 | 1.205 | 0.228 | 0.999 |
| | Right Hippocampus | 0.233 | 0.189 | 1.232 | 0.218 | 0.999 |
| | WMH | 0.020 | 0.171 | 0.115 | 0.909 | 0.999 |
| Bioidentical & Synthetic | GM BAG | -0.008 | 0.107 | -0.075 | 0.940 | 0.999 |
| | WM BAG | 0.056 | 0.108 | 0.519 | 0.604 | 0.999 |
| | Left Hippocampus | -0.061 | 0.101 | -0.604 | 0.546 | 0.999 |
| | Right Hippocampus | -0.083 | 0.102 | -0.817 | 0.414 | 0.999 |
| | WMH | 0.037 | 0.092 | 0.407 | 0.684 | 0.999 |
| **Estrogens + Progestins, active ingredient** | | | | | | |
| estradiol hemihydrate & norethisterone acetate | GM BAG | -0.058 | 0.142 | -0.405 | 0.685 | 0.999 |
| | WM BAG | -0.001 | 0.143 | -0.006 | 0.995 | 0.999 |
| | Left Hippocampus | 0.042 | 0.135 | 0.310 | 0.757 | 0.999 |
| | Right Hippocampus | -0.013 | 0.135 | -0.094 | 0.925 | 0.999 |
| | WMH | 0.026 | 0.122 | 0.213 | 0.832 | 0.999 |
| estradiol hemihydrate & dydrogesterone | GM BAG | -0.362 | 0.276 | -1.315 | 0.188 | 0.999 |
| | WM BAG | -0.121 | 0.277 | -0.438 | 0.661 | 0.999 |
| | Left Hippocampus | 0.024 | 0.261 | 0.090 | 0.928 | 0.999 |

| | | | | | | |
|---|---|---|---|---|---|---|
| | Right Hippocampus | 0.387 | 0.261 | 1.480 | 0.139 | 0.999 |
| | WMH | -0.105 | 0.236 | -0.447 | 0.655 | 0.999 |
| estradiol hemihydrate & norethisterone | GM BAG | 0.287 | 0.287 | 1.001 | 0.317 | 0.999 |
| | WM BAG | 0.193 | 0.288 | 0.669 | 0.504 | 0.999 |
| | Left Hippocampus | -0.215 | 0.272 | -0.791 | 0.429 | 0.999 |
| | Right Hippocampus | -0.286 | 0.272 | -1.052 | 0.293 | 0.999 |
| | WMH | 0.369 | 0.245 | 1.506 | 0.132 | 0.999 |
| CEE & norgestrel | GM BAG | -0.116 | 0.241 | -0.480 | 0.632 | 0.999 |
| | WM BAG | -0.210 | 0.242 | -0.866 | 0.387 | 0.999 |
| | Left Hippocampus | 0.068 | 0.228 | 0.296 | 0.767 | 0.999 |
| | Right Hippocampus | -0.051 | 0.229 | -0.221 | 0.825 | 0.999 |
| | WMH | 0.098 | 0.206 | 0.474 | 0.635 | 0.999 |
| CEE & medroxyprogesterone acetate | GM BAG | -0.267 | 0.351 | -0.761 | 0.447 | 0.999 |
| | WM BAG | 0.403 | 0.353 | 1.141 | 0.254 | 0.999 |
| | Left Hippocampus | 0.558 | 0.333 | 1.678 | 0.093 | 0.999 |
| | Right Hippocampus | 0.830 | 0.333 | 2.493 | **0.013** | 0.847 |
| | WMH | -0.147 | 0.300 | -0.490 | 0.624 | 0.999 |
| tibolone | GM BAG | -0.418 | 0.276 | -1.514 | 0.130 | 0.999 |
| | WM BAG | 0.036 | 0.277 | 0.129 | 0.898 | 0.999 |
| | Left Hippocampus | 0.016 | 0.261 | 0.063 | 0.950 | 0.999 |
| | Right Hippocampus | 0.153 | 0.262 | 0.584 | 0.560 | 0.999 |
| | WMH | -0.156 | 0.245 | -0.637 | 0.524 | 0.999 |
| Mixed | GM BAG | 0.116 | 0.083 | 1.408 | 0.159 | 0.999 |
| | WM BAG | 0.172 | 0.083 | 2.069 | **0.039** | 0.986 |
| | Left Hippocampus | 0.035 | 0.078 | 0.451 | 0.652 | 0.999 |
| | Right Hippocampus | -0.030 | 0.078 | -0.383 | 0.702 | 0.999 |
| | WMH | 0.064 | 0.072 | 0.881 | 0.378 | 0.999 |
| **Estrogens + Progestins, Progestin Generation** | | | | | | |
| 1stGen | GM BAG | 0.035 | 0.097 | 0.364 | 0.716 | 0.999 |
| | WM BAG | 0.136 | 0.097 | 1.393 | 0.164 | 0.999 |
| | Left Hippocampus | 0.013 | 0.092 | 0.144 | 0.886 | 0.999 |
| | Right Hippocampus | 0.000 | 0.092 | -0.001 | 0.999 | 0.999 |
| | WMH | 0.061 | 0.084 | 0.722 | 0.470 | 0.999 |
| 2ndGen | GM BAG | -0.064 | 0.162 | -0.393 | 0.695 | 0.999 |
| | WM BAG | -0.011 | 0.162 | -0.068 | 0.946 | 0.999 |
| | Left Hippocampus | 0.044 | 0.153 | 0.288 | 0.773 | 0.999 |
| | Right Hippocampus | 0.084 | 0.153 | 0.549 | 0.583 | 0.999 |
| | WMH | 0.082 | 0.139 | 0.595 | 0.552 | 0.999 |
| **Estrogens + Progestins, Dosage (mg)** | | | | | | |
| Estrogens | GM BAG | -0.075 | 0.059 | -1.264 | 0.207 | 0.999 |
| | WM BAG | -0.003 | 0.059 | -0.044 | 0.965 | 0.999 |
| | Left Hippocampus | -0.074 | 0.057 | -1.293 | 0.197 | 0.999 |
| | Right Hippocampus | -0.009 | 0.057 | -0.158 | 0.875 | 0.999 |
| | WMH | 0.047 | 0.053 | 0.887 | 0.376 | 0.999 |
| Progestins | GM BAG | 0.064 | 0.057 | 1.132 | 0.258 | 0.999 |
| | WM BAG | 0.055 | 0.057 | 0.965 | 0.335 | 0.999 |
| | Left Hippocampus | 0.063 | 0.055 | 1.160 | 0.247 | 0.999 |
| | Right Hippocampus | 0.041 | 0.054 | 0.765 | 0.445 | 0.999 |
| | WMH | 0.071 | 0.050 | 1.416 | 0.158 | 0.999 |
| **Estrogens + Progestins, Duration of Use (weeks)** | | | | | | |
| Estrogens | GM BAG | 0.161 | 0.081 | 1.984 | **0.048** | 0.986 |
| | WM BAG | -0.018 | 0.083 | -0.214 | 0.831 | 0.999 |
| | Left Hippocampus | -0.170 | 0.077 | -2.204 | **0.028** | 0.986 |
| | Right Hippocampus | -0.183 | 0.075 | -2.428 | **0.016** | 0.847 |
| | WMH | -0.105 | 0.074 | -1.432 | 0.153 | 0.999 |

| | | | | | | |
|---|---|---|---|---|---|---|
| Progestins | GM BAG | -0.042 | 0.081 | -0.518 | 0.605 | 0.999 |
| | WM BAG | 0.087 | 0.083 | 1.042 | 0.298 | 0.999 |
| | Left Hippocampus | -0.025 | 0.077 | -0.328 | 0.743 | 0.999 |
| | Right Hippocampus | 0.028 | 0.075 | 0.377 | 0.707 | 0.999 |
| | WMH | 0.089 | 0.073 | 1.218 | 0.225 | 0.999 |

Significant results are highlighted in bold. False discovery rate (FDR) correction was applied across all brain measures and MHT variables listed in this table. Abbreviations: MRI = magnetic resonance imaging, S.E. = standard error, GM = grey matter, BAG = brain age gap, WM = white matter, WMH = white matter hyperintensity, CEE = conjugated equine estrogen, Gen = generation.

**Table S15| Associations between menopausal hormone therapy (MHT)-related variables and brain measures in the prescription MHT sample, with age, education, and menopause-status matched never-users.**

| MHT Variable | MRI Measure | beta | S.E. | t-value | p-value | pFDR-value |
|---|---|---|---|---|---|---|
| **MHT formulation** | | | | | | |
| Estrogens-only | GM BAG | -0.071 | 0.080 | -0.889 | 0.374 | 0.986 |
| | WM BAG | -0.009 | 0.080 | -0.112 | 0.911 | 0.986 |
| | Left Hippocampus | 0.008 | 0.075 | 0.106 | 0.916 | 0.986 |
| | Right Hippocampus | -0.006 | 0.075 | -0.087 | 0.931 | 0.986 |
| | WMH | 0.067 | 0.071 | 0.950 | 0.342 | 0.986 |
| Estrogens+Progestin | GM BAG | -0.027 | 0.071 | -0.377 | 0.706 | 0.986 |
| | WM BAG | 0.088 | 0.071 | 1.233 | 0.218 | 0.986 |
| | Left Hippocampus | 0.051 | 0.067 | 0.764 | 0.445 | 0.986 |
| | Right Hippocampus | 0.019 | 0.067 | 0.280 | 0.779 | 0.986 |
| | WMH | 0.099 | 0.063 | 1.575 | 0.115 | 0.986 |
| **Route of Administration** | | | | | | |
| oral | GM BAG | -0.083 | 0.078 | -1.060 | 0.289 | 0.986 |
| | WM BAG | 0.068 | 0.079 | 0.869 | 0.385 | 0.986 |
| | Left Hippocampus | 0.036 | 0.074 | 0.485 | 0.628 | 0.986 |
| | Right Hippocampus | 0.034 | 0.074 | 0.459 | 0.646 | 0.986 |
| | WMH | 0.178 | 0.069 | 2.586 | **0.010** | 0.986 |
| transdermal | GM BAG | -0.247 | 0.132 | -1.868 | 0.062 | 0.986 |
| | WM BAG | -0.082 | 0.132 | -0.621 | 0.535 | 0.986 |
| | Left Hippocampus | 0.032 | 0.125 | 0.254 | 0.800 | 0.986 |
| | Right Hippocampus | -0.015 | 0.124 | -0.121 | 0.904 | 0.986 |
| | WMH | -0.034 | 0.117 | -0.290 | 0.772 | 0.986 |
| vaginal | GM BAG | 0.101 | 0.105 | 0.958 | 0.338 | 0.986 |
| | WM BAG | 0.104 | 0.105 | 0.987 | 0.324 | 0.986 |
| | Left Hippocampus | 0.010 | 0.100 | 0.098 | 0.922 | 0.986 |
| | Right Hippocampus | 0.026 | 0.099 | 0.264 | 0.792 | 0.986 |
| | WMH | 0.118 | 0.094 | 1.251 | 0.211 | 0.986 |
| injection | GM BAG | 0.377 | 0.336 | 1.121 | 0.262 | 0.986 |
| | WM BAG | 0.159 | 0.337 | 0.472 | 0.637 | 0.986 |
| | Left Hippocampus | -0.191 | 0.319 | -0.598 | 0.550 | 0.986 |
| | Right Hippocampus | 0.138 | 0.316 | 0.437 | 0.662 | 0.986 |
| | WMH | -0.018 | 0.291 | -0.060 | 0.952 | 0.986 |
| mixed | GM BAG | -0.028 | 0.100 | -0.282 | 0.778 | 0.986 |
| | WM BAG | 0.017 | 0.100 | 0.172 | 0.864 | 0.986 |
| | Left Hippocampus | 0.066 | 0.095 | 0.699 | 0.485 | 0.986 |
| | Right Hippocampus | -0.054 | 0.094 | -0.570 | 0.569 | 0.986 |
| | WMH | -0.050 | 0.088 | -0.563 | 0.574 | 0.986 |
| **Estrogen-only Forms** | | | | | | |
| Bioidentical | GM BAG | -0.052 | 0.087 | -0.594 | 0.553 | 0.986 |
| | WM BAG | 0.002 | 0.087 | 0.018 | 0.986 | 0.986 |
| | Left Hippocampus | 0.025 | 0.082 | 0.304 | 0.761 | 0.986 |
| | Right Hippocampus | 0.018 | 0.081 | 0.219 | 0.827 | 0.986 |
| | WMH | 0.056 | 0.077 | 0.726 | 0.468 | 0.986 |
| Synthetic | GM BAG | -0.030 | 0.218 | -0.138 | 0.890 | 0.986 |
| | WM BAG | -0.056 | 0.218 | -0.257 | 0.797 | 0.986 |
| | Left Hippocampus | -0.017 | 0.205 | -0.085 | 0.932 | 0.986 |
| | Right Hippocampus | 0.026 | 0.203 | 0.126 | 0.900 | 0.986 |
| | WMH | 0.133 | 0.187 | 0.711 | 0.477 | 0.986 |
| **Estrogen-only, active ingredient** | | | | | | |
| estradiol | GM BAG | -0.284 | 0.177 | -1.606 | 0.109 | 0.986 |
| | WM BAG | -0.186 | 0.177 | -1.051 | 0.294 | 0.986 |
| | Left Hippocampus | 0.034 | 0.166 | 0.204 | 0.838 | 0.986 |
| | Right Hippocampus | 0.068 | 0.164 | 0.416 | 0.677 | 0.986 |
| | WMH | 0.003 | 0.154 | 0.018 | 0.986 | 0.986 |

| | | | | | | |
|---|---|---|---|---|---|---|
| estradiol hemihydrate | GM BAG | 0.005 | 0.095 | 0.050 | 0.960 | 0.986 |
| | WM BAG | 0.047 | 0.095 | 0.492 | 0.623 | 0.986 |
| | Left Hippocampus | 0.022 | 0.089 | 0.251 | 0.802 | 0.986 |
| | Right Hippocampus | 0.005 | 0.088 | 0.061 | 0.952 | 0.986 |
| | WMH | 0.069 | 0.084 | 0.825 | 0.410 | 0.986 |
| estradiol valerate | GM BAG | 0.409 | 0.502 | 0.814 | 0.416 | 0.986 |
| | WM BAG | 0.191 | 0.503 | 0.380 | 0.704 | 0.986 |
| | Left Hippocampus | 0.062 | 0.471 | 0.131 | 0.896 | 0.986 |
| | Right Hippocampus | 0.427 | 0.465 | 0.920 | 0.358 | 0.986 |
| | WMH | 0.214 | 0.429 | 0.500 | 0.617 | 0.986 |
| CEE | GM BAG | -0.129 | 0.241 | -0.537 | 0.591 | 0.986 |
| | WM BAG | -0.109 | 0.241 | -0.453 | 0.651 | 0.986 |
| | Left Hippocampus | -0.032 | 0.226 | -0.144 | 0.886 | 0.986 |
| | Right Hippocampus | -0.061 | 0.222 | -0.273 | 0.785 | 0.986 |
| | WMH | 0.115 | 0.206 | 0.558 | 0.577 | 0.986 |
| Mixed | GM BAG | -0.255 | 0.198 | -1.292 | 0.197 | 0.986 |
| | WM BAG | -0.047 | 0.198 | -0.236 | 0.813 | 0.986 |
| | Left Hippocampus | -0.066 | 0.185 | -0.354 | 0.724 | 0.986 |
| | Right Hippocampus | -0.168 | 0.183 | -0.917 | 0.360 | 0.986 |
| | WMH | 0.075 | 0.172 | 0.437 | 0.662 | 0.986 |
| **Estrogens + Progestins Form** | | | | | | |
| Bioidentical | GM BAG | -0.148 | 0.247 | -0.598 | 0.550 | 0.986 |
| | WM BAG | -0.023 | 0.261 | -0.087 | 0.931 | 0.986 |
| | Left Hippocampus | 0.143 | 0.241 | 0.594 | 0.553 | 0.986 |
| | Right Hippocampus | 0.317 | 0.243 | 1.303 | 0.193 | 0.986 |
| | WMH | 0.080 | 0.224 | 0.356 | 0.722 | 0.986 |
| Synthetic | GM BAG | -0.050 | 0.187 | -0.270 | 0.787 | 0.986 |
| | WM BAG | 0.149 | 0.197 | 0.753 | 0.452 | 0.986 |
| | Left Hippocampus | 0.220 | 0.182 | 1.208 | 0.228 | 0.986 |
| | Right Hippocampus | 0.235 | 0.184 | 1.277 | 0.202 | 0.986 |
| | WMH | 0.177 | 0.170 | 1.046 | 0.296 | 0.986 |
| Bioidentical & Synthetic | GM BAG | -0.027 | 0.111 | -0.247 | 0.805 | 0.986 |
| | WM BAG | 0.023 | 0.117 | 0.192 | 0.848 | 0.986 |
| | Left Hippocampus | -0.052 | 0.108 | -0.477 | 0.634 | 0.986 |
| | Right Hippocampus | -0.112 | 0.110 | -1.019 | 0.309 | 0.986 |
| | WMH | 0.120 | 0.102 | 1.177 | 0.240 | 0.986 |
| **Estrogens + Progestins, active ingredient** | | | | | | |
| estradiol hemihydrate & norethisterone acetate | GM BAG | -0.099 | 0.146 | -0.679 | 0.497 | 0.986 |
| | WM BAG | -0.044 | 0.148 | -0.299 | 0.765 | 0.986 |
| | Left Hippocampus | 0.095 | 0.140 | 0.677 | 0.499 | 0.986 |
| | Right Hippocampus | 0.003 | 0.139 | 0.018 | 0.986 | 0.986 |
| | WMH | 0.112 | 0.129 | 0.870 | 0.385 | 0.986 |
| estradiol hemihydrate & dydrogesterone | GM BAG | -0.224 | 0.269 | -0.833 | 0.405 | 0.986 |
| | WM BAG | -0.120 | 0.272 | -0.439 | 0.660 | 0.986 |
| | Left Hippocampus | 0.086 | 0.257 | 0.336 | 0.737 | 0.986 |
| | Right Hippocampus | 0.320 | 0.255 | 1.251 | 0.211 | 0.986 |
| | WMH | 0.042 | 0.235 | 0.178 | 0.859 | 0.986 |
| estradiol hemihydrate & norethisterone | GM BAG | 0.222 | 0.280 | 0.794 | 0.428 | 0.986 |
| | WM BAG | 0.099 | 0.283 | 0.351 | 0.726 | 0.986 |
| | Left Hippocampus | -0.308 | 0.267 | -1.152 | 0.250 | 0.986 |
| | Right Hippocampus | -0.426 | 0.265 | -1.605 | 0.109 | 0.986 |
| | WMH | 0.407 | 0.244 | 1.667 | 0.096 | 0.986 |
| CEE & norgestrel | GM BAG | -0.098 | 0.234 | -0.418 | 0.676 | 0.986 |
| | WM BAG | -0.102 | 0.236 | -0.434 | 0.664 | 0.986 |
| | Left Hippocampus | 0.212 | 0.223 | 0.951 | 0.342 | 0.986 |
| | Right Hippocampus | 0.110 | 0.222 | 0.496 | 0.620 | 0.986 |

| | | | | | | |
|---|---|---|---|---|---|---|
| | WMH | 0.166 | 0.204 | 0.816 | 0.415 | 0.986 |
| CEE & medroxyprogesterone acetate | GM BAG | 0.042 | 0.318 | 0.134 | 0.894 | 0.986 |
| | WM BAG | 0.566 | 0.321 | 1.765 | 0.078 | 0.986 |
| | Left Hippocampus | 0.198 | 0.303 | 0.652 | 0.515 | 0.986 |
| | Right Hippocampus | 0.440 | 0.301 | 1.461 | 0.145 | 0.986 |
| | WMH | 0.167 | 0.277 | 0.601 | 0.548 | 0.986 |
| tibolone | GM BAG | -0.446 | 0.282 | -1.582 | 0.114 | 0.986 |
| | WM BAG | -0.069 | 0.285 | -0.241 | 0.810 | 0.986 |
| | Left Hippocampus | 0.135 | 0.269 | 0.500 | 0.617 | 0.986 |
| | Right Hippocampus | 0.264 | 0.267 | 0.988 | 0.323 | 0.986 |
| | WMH | -0.154 | 0.255 | -0.602 | 0.547 | 0.986 |
| Mixed | GM BAG | 0.032 | 0.089 | 0.355 | 0.722 | 0.986 |
| | WM BAG | 0.136 | 0.090 | 1.513 | 0.131 | 0.986 |
| | Left Hippocampus | 0.058 | 0.085 | 0.681 | 0.496 | 0.986 |
| | Right Hippocampus | -0.004 | 0.084 | -0.042 | 0.966 | 0.986 |
| | WMH | 0.084 | 0.079 | 1.063 | 0.288 | 0.986 |
| **Estrogens + Progestins, Progestin Generation** | | | | | | |
| 1stGen | GM BAG | 0.019 | 0.104 | 0.179 | 0.858 | 0.986 |
| | WM BAG | 0.101 | 0.104 | 0.977 | 0.329 | 0.986 |
| | Left Hippocampus | 0.007 | 0.097 | 0.067 | 0.946 | 0.986 |
| | Right Hippocampus | -0.029 | 0.096 | -0.299 | 0.765 | 0.986 |
| | WMH | 0.130 | 0.093 | 1.391 | 0.165 | 0.986 |
| 2ndGen | GM BAG | -0.035 | 0.162 | -0.216 | 0.829 | 0.986 |
| | WM BAG | 0.021 | 0.162 | 0.128 | 0.898 | 0.986 |
| | Left Hippocampus | 0.148 | 0.152 | 0.973 | 0.331 | 0.986 |
| | Right Hippocampus | 0.142 | 0.151 | 0.939 | 0.348 | 0.986 |
| | WMH | 0.164 | 0.144 | 1.138 | 0.255 | 0.986 |

Significant results are highlighted in bold. False discovery rate (FDR) correction was applied across all brain measures and MHT variables listed in this table. Abbreviations: MRI = magnetic resonance imaging, S.E. = standard error, GM = grey matter, BAG = brain age gap, WM = white matter, WMH = white matter hyperintensity, CEE = conjugated equine estrogen, Gen = generation.
